# Integrated single-cell and proteomic profiling of pancreatic neoplasia identifies tissue markers that outperform established diagnostic markers in distinguishing cancer from precursor lesions

**DOI:** 10.64898/2026.10.02.26364030

**Authors:** Thoa T. Tran, Phuong-Uyen Nguyen-Hoang, Johannes F. Fahrmann, Duy Truong Pham, Huy Bui, Loan Bui, C. Max Schmidt

## Abstract

Pancreatic ductal adenocarcinoma (PDAC) arises within an extensively remodeled microenvironment, yet tissue markers that distinguish invasive carcinoma from precursor lesions remain limited. Integrating four single-cell RNA-sequencing atlases comprising 324,662 cells from 26 patients, we identified cancer-associated fibroblast expansion, accumulation of C1QA+ immunoregulatory macrophages, and depletion of a dendritic-cell compartment expressing plasmacytoid markers. Fibroblasts were enriched for extracellular-matrix and PI3K–AKT programs and macrophages for an antigen-presentation program. We then examined the protein-level correlates of these populations in a tissue proteomic reference framework of 356 samples spanning non-diseased pancreas, normal-adjacent tissue, low- and high-grade intraductal papillary mucinous neoplasms (IPMNs), IPMN-associated invasive carcinoma, and PDAC, prioritizing proteins that changed monotonically across the six histopathologic reference groups or marked tumor-enriched cell types. Despite depletion of plasmacytoid dendritic cells, the principal source of type I interferon, the interferon-stimulated protein ISG15, expressed by both fibroblasts and macrophages, increased monotonically toward PDAC. In exploratory cross-validated benchmarking of PDAC versus low- and high-grade IPMN, excluding IPMN-associated invasive carcinoma (145 versus 56 samples), CATB+EPIPL achieved an AUC of 0.98, with a sensitivity of 92% at an achieved specificity of 92%, exceeding protein-abundance models of diagnostic IHC-associated markers (S100P+IMP3+maspin+MSLN, 0.79) and refitted published IPMN-progression combinations (≤0.91). Against non-diseased pancreas, candidate and comparator panels performed similarly (AUC 0.99–1.00). These biologically informed tissue protein panels warrant independent validation, including immunohistochemical evaluation of EUS-guided fine-needle biopsy specimens and measurement in blood.

## Introduction

Pancreatic ductal adenocarcinoma (PDAC) remains one of the most lethal human malignancies, characterized by diagnosis at an advanced stage, extensive stromal desmoplasia, profound immune suppression, and limited durable responses to current therapies^1,2^. Although tumor-cell-intrinsic alterations such as KRAS activation, TP53 loss, and cell-cycle dysregulation are central to PDAC biology, disease progression is increasingly understood as a systemic, multicellular process rather than a tumor-cell-autonomous one^2,3^. Recent success of tumor-cell-intrinsic RAS(ON) inhibition in metastatic PDAC^4^ highlight the need to define complementary tumor microenvironment (TME) programs that may independently shape disease progression and therapeutic response. Current detection strategies rely predominantly on single-timepoint, single-modality case-control comparisons, an approach whose predictive performance remains limited even with multi-marker ensemble ^5,6^ and that is poorly suited to a disease that develops through a progressive, multi-stage remodeling of both the neoplastic epithelium and its surrounding microenvironment: a biomarker identified from one pairwise stage contrast may not generalize across the full pancreatic neoplasia spectrum, and imaging-based surveillance alone cannot resolve the underlying molecular heterogeneity that determines which lesions will ultimately progress to malignancy^7,8^. Capturing the true pattern of neoplastic progression, and the specific biological advantage it offers for early and specific detection, therefore requires integrating single-cell resolution of the cellular and signaling programs that drive disease evolution with pathway-level modeling and tissue-level proteomic profiling capable of validating candidate biomarkers across the full disease spectrum, an integrative multi-omics strategy already shown to resolve clinically relevant subtypes and therapeutic vulnerabilities in PDAC at the bulk tissue level^9–12^ single-cell transcriptomic data alone cannot establish which candidate molecules are detectable in patient tissue or serum, and bulk proteomic data alone cannot resolve which cell type or signaling pathway produces a given protein signal.

The PDAC tumor microenvironment (TME) is composed of neoplastic epithelial cells, cancer-associated fibroblasts (CAFs), tumor-associated macrophages (TAMs), T cells, dendritic cells, endothelial cells, and other stromal and immune populations that interact through extracellular matrix remodeling, inflammatory signaling, immune checkpoint regulation, angiogenesis, and metabolic adaptation^13–18^, contributing to local invasion, immune evasion, invasion of nerves and vessels of nerves and vessels, and therapeutic resistance^9,19,20^. CAFs generate extracellular-matrix-rich stromal scaffolds that regulate tumor stiffness and growth factors and have been resolved by single-cell profiling into functionally distinct subtypes, including an antigen-presenting CAF state^9,15,21–23^ while TAMs contribute to inflammation, tissue repair, angiogenesis, and suppression of cytotoxic immunity^13,18,24–28^.

Large-scale single-cell and spatial profiling efforts have progressively resolved this complexity: early surveys established substantial intratumoral heterogeneity and neoplastic-cell progression states in PDAC^9,12,15,26,27,29^ spatially confined sub-tumor microenvironments were subsequently shown to organize this heterogeneity into discrete ecological niches^9,12,15,26,27,29^ with high-resolution spatial mapping further resolving the tissue architecture in which these niches are embedded^15,16,23^ Chen and colleagues used integrated single-cell and spatial transcriptomics to identify distinct cellular subtypes mediating periinvasion of nerves and vessels; Caronni and colleagues defined a pathogenic interleukin-1-beta-producing macrophage population that fuels inflammation and immunosuppression^30^, consistent with reports that SPP1-expressing macrophages define a conserved cellular program controlling tumor progression across cancer types^25^. Zhang and colleagues characterized an immunosuppressive tumor microenvironment specific to PDAC liver metastasis^17^; Zheng and colleagues integrated single-cell and bulk RNA sequencing to derive a prognostic T-cell marker gene score^16^; PDAC tumor-associated macrophages have been shown to originate substantially from embryonic, tissue-resident precursors^31^ and to display phenotypic and spatial heterogeneity independently associated with clinical outcome^32^; Kanu and colleagues established a multisite single-cell immune-profiling platform for premetastatic PDAC, showing that immune states differ across the tumor, blood, and premetastatic liver^33^; and Tang and colleagues assembled a large-scale, cell-type-resolved single-nucleus transcriptomic atlas of PDAC linked to patient survival^9^. Building on this single-cell foundation, Wang and colleagues, from the Hruban and Zhang laboratories at Johns Hopkins University, reported a comprehensive proteomic and glycoproteomic analysis of IPMNs and PDACs, identifying tumor-progression pathways enriched with disease grade and validating candidate markers including PLOD3, IRS2, LGALS9, and Trop-2^29^, each with independent mechanistic support in the broader cancer literature^34–38^, with complementary spatial proteomic work further resolving protein-expression changes across normal-duct, IPMN, and PDAC tissue architecture^10^. Despite these advances, most single-cell PDAC studies remain limited by small cohort size and dataset-specific variation, and these single-cell and spatial characterizations of the TME have generally been reported separately from tissue-level proteomic biomarker development; prior proteomic panels, in turn, have focused predominantly on proteins upregulated in disease, leaving open whether single-cell-defined cellular programs, reproduced across independent cohorts, can directly guide which circulating or tissue proteins are most likely to discriminate disease stage and serve as therapeutic targets.

Here, we integrated four independent single-cell RNA-sequencing atlases of PDAC^15,16,30,33^ to characterize reproducible stromal and immune remodeling patterns, with particular emphasis on cancer-associated fibroblasts, tumor-associated macrophages, and plasmacytoid dendritic cells. We then examined these findings alongside an independent tissue proteomic cohort spanning non-diseased pancreas, normal-adjacent tissue, low-grade and high-grade IPMN, IPMN-associated invasive carcinoma, and PDAC. We evaluated proteins with increased or decreased abundance, individually and in combination, to identify biologically informed candidates for PDAC detection, assess their behavior across clinically relevant pancreatic lesions, and explore their potential relevance to therapeutic targeting.

## Materials and Methods

### 1. Data Collection

Publicly available single-cell RNA sequencing (scRNA-seq) datasets of treatment-naïve pancreatic ductal adenocarcinoma (PDAC) were obtained from the Gene Expression Omnibus (GEO), including GSE278694^15^, GSE212966^16^, GSE217845^30^, and GSE300304^33^. Clinical information, sample annotations, and processed gene-expression matrices were retrieved for all available samples.

To ensure consistency across cohorts and enable stage-based comparisons, only primary treatment-naïve PDAC tumor samples with available clinical stage information were included in the final integrated analysis. After sample selection and quality control, the final cohort consisted of 26 PDAC patients representing clinical stages I–IV, including 9 stage I tumors, 8 stage II tumors, 8 stage III tumors, and 1 stage IV tumor (Figure 1A). The integrated dataset comprised 324,662 high-quality single cells.

**Figure 1.**
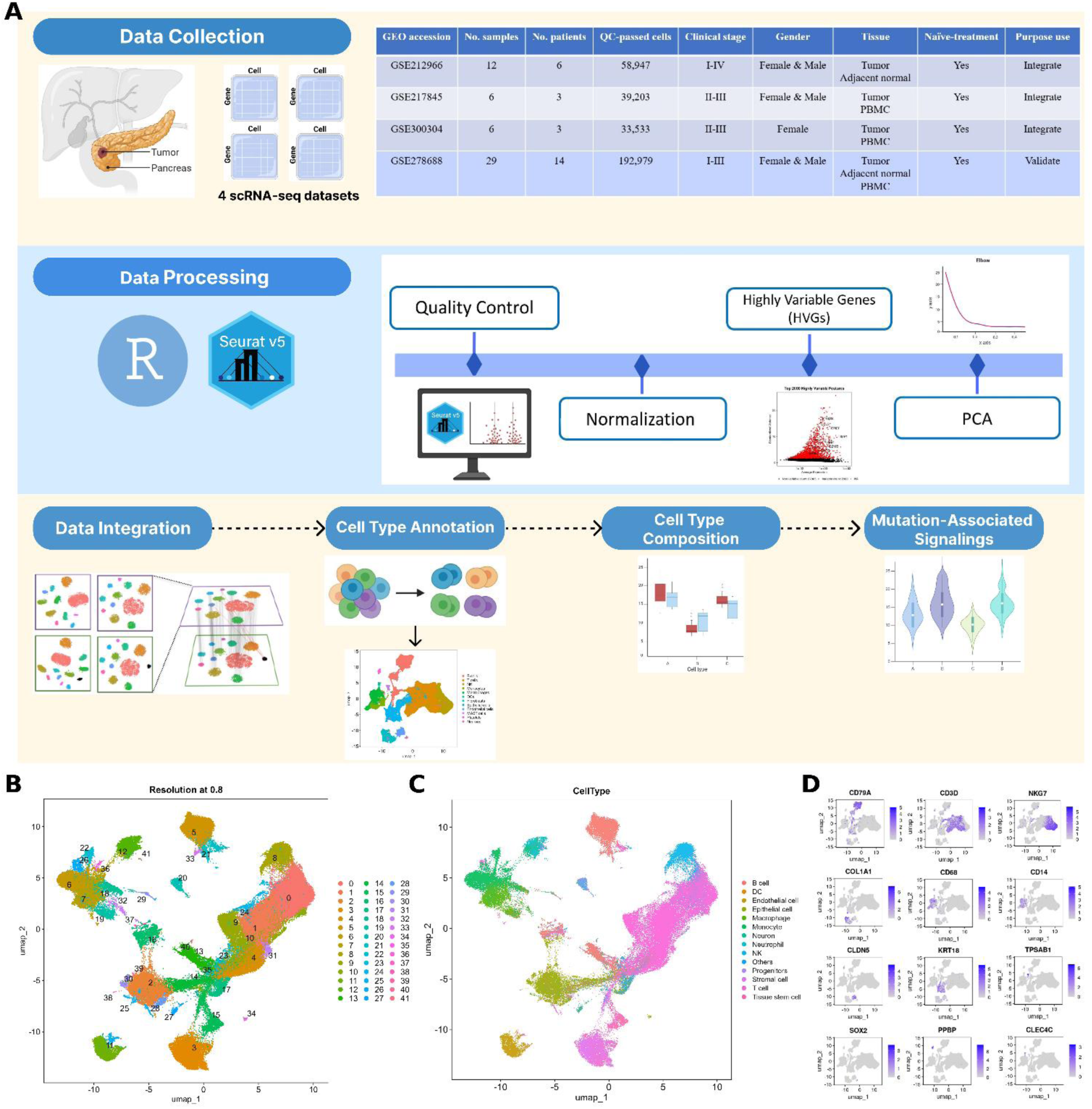
Integrated single-cell atlas of treatment-naive PDAC. (A) Study workflow (schematic overview; final cohort composition and cell numbers are reported in Methods). Four publicly available scRNA-seq datasets were obtained from GEO (the table lists the number of samples, patients, and QC-passed cells, clinical stage, sex, tissue source, treatment status, and use in the analysis), processed in R with Seurat v5 (quality control, normalization, selection of the top 2,000 highly variable genes, and principal component analysis), integrated with Harmony, and used for cell-type annotation, composition, and pathway-signaling analyses. (B) UMAP of the integrated atlas (324,662 cells from 26 patients) colored by unsupervised graph-based cluster (resolution 0.8). (C) UMAP colored by annotated cell type. (D) Feature plots of canonical lineage markers: CD79A (B cells), CD3D (T cells), NKG7 (NK cells), COL1A1 (fibroblasts), CD68 (macrophages), CD14 (monocytes), CLDN5 (endothelial cells), KRT18 (epithelial cells), TPSAB1 (mast cells), SOX2 (neurons), PPBP (platelets), and CLEC4C (plasmacytoid dendritic cells). Color scale indicates log-normalized expression.

The final cohort included samples from five independent studies: GSE278694 (14 patients)^15^, GSE212966 (6 patients)^16^, GSE217845 (3 patients)^30^, GSE300304 (3 patients)^33^, and the validation cohort GSE197177 (4 patients)^17^. All patients were treatment-naïve at the time of tissue collection, minimizing treatment-induced transcriptional alterations and enabling characterization of the native tumor microenvironment during PDAC progression.

Raw count matrices and associated metadata were processed using a unified analytical workflow. Dataset-specific quality-control procedures were applied to remove low-quality cells, doublets, and potential technical artifacts before integration. Following normalization and batch correction, cells from all cohorts were combined to generate an integrated PDAC single-cell atlas. This atlas was subsequently used for cell-type annotation, differential gene expression analysis, pathway enrichment analysis, protein-protein interaction network analysis, and characterization of stromal and immune remodeling during PDAC progression.

### 2. scRNA-seq Data Quality Control

Preprocessed gene expression matrices and accompanying cell annotations were analyzed using the Seurat package (v5.3.1)^39^ in R (v4.3.2). Quality control (QC) was performed independently for each dataset prior to integration to remove low-quality cells and technical artifacts. Three commonly used metrics were evaluated for each cell: the number of detected genes (nFeature_RNA), total unique molecular identifiers (nCount_RNA), and the percentage of mitochondrial gene expression (percent.mt). For GSE278694, genes detected in fewer than three cells were excluded. Cells with fewer than 200 detected genes, more than 5,000 detected genes, greater than 20,000 UMIs, or more than 10% mitochondrial transcripts were removed. The original dataset contained 30,543 genes and 199,184 cells; after quality control, 192,979 high-quality cells and 29,065 genes were retained (Supplementary Figure S1). For GSE212966, genes expressed in fewer than three cells were excluded. Cells with fewer than 200 detected genes, more than 8,000 detected genes, greater than 60,000 UMIs, or more than 20% mitochondrial transcripts were removed. Following filtering, 32,431 high-quality cells were retained from an initial 69,105 cells, while 25,579 genes remained for downstream analysis (Supplementary Figure S2). For GSE217845, genes detected in fewer than three cells were removed. Cells with fewer than 200 detected genes, more than 5,000 detected genes, greater than 50,000 UMIs, or more than 25% mitochondrial transcripts were excluded. After quality control, 47,635 high-quality cells and 24,172 genes were retained from the original 60,060 cells (Supplementary Figure S3). For GSE300304, genes expressed in fewer than three cells were filtered out. Cells with fewer than 200 detected genes, more than 5,000 detected genes, greater than 25,000 UMIs, or more than 20% mitochondrial transcripts were removed. Following filtering, 33,533 high-quality cells and 23,832 genes were retained from the original 34,982 cells (Supplementary Figure S4).

After dataset-specific quality control, eligible treatment-naïve PDAC tumor samples with available clinical stage information were selected for integration. The final integrated cohort consisted of 26 PDAC patients spanning stages I–IV and included 324,662 high-quality cells for downstream analyses.

### 3. scRNA-seq Data Preprocessing, Integration, and Cluster Identification

Following quality control and selection of eligible treatment naïve PDAC tumor samples, a total of 324,662 high-quality cells were retained for downstream analysis. Gene expression matrices from all datasets were processed using the Seurat package (v5.3.1). Expression values were normalized using the “NormalizeData” function to correct for differences in sequencing depth and library size across cells. Highly variable genes (HVGs) were subsequently identified using the “FindVariableFeatures” function, and the top 2,000 HVGs were selected for downstream analyses (Supplementary Figure S5A). Gene expression values were then scaled using the “ScaleData” function to center and standardize expression levels across cells.

To capture the major sources of biological variation, principal component analysis (PCA) was performed using the selected highly variable genes. PCA reduces the dimensionality of the dataset while preserving the most informative transcriptional patterns. Examination of the elbow plot indicated that the first 40 principal components captured most of the biologically meaningful variation and were therefore retained for downstream analyses (Supplementary Figure S5B).

Because cells originated from multiple independent studies and patients, potential batch effects were corrected using the Harmony algorithm (v1.2.4)^40^. Harmony integrates datasets by aligning shared biological signals while minimizing study-specific technical variation, thereby enabling robust cross-cohort analysis while preserving biologically relevant cellular heterogeneity (Supplementary Figure S6).

Unsupervised graph-based clustering was subsequently performed using Harmony-corrected embeddings. A shared nearest-neighbor graph was constructed using the first 40 Harmony dimensions with the “FindNeighbors” function, and cell clusters were identified using the “FindClusters” function with a resolution parameter of 0.8. This analysis identified 42 transcriptionally distinct cell clusters. Cellular relationships were visualized using Uniform Manifold Approximation and Projection (UMAP), which projected high-dimensional transcriptional profiles into a two-dimensional space while preserving local and global cellular structure.

The resulting integrated atlas served as the foundation for downstream cell-type annotation, differential expression analysis, pathway enrichment analysis, protein–protein interaction network analysis, and characterization of stromal and immune remodeling associated with PDAC progression. Subsequent analyses focused on fibroblast, macrophage, dendritic-cell, endothelial-cell, and lymphoid compartments to investigate multicellular stromal–immune remodeling associated with PDAC progression.

### 4. Differential Gene Expression Analysis and Cell-Type Annotation

Differentially expressed genes (DEGs) for each cluster were identified using the Wilcoxon rank-sum test implemented in the Seurat “FindAllMarkers” function. Marker genes were required to be expressed in at least 50% of cells within a cluster and exhibit a minimum average log2 fold change of 0.25 relative to all remaining cells. The complete marker gene lists for each cell type are provided in Supplementary Table S2. The resulting DEG lists were used to define cluster-specific transcriptional signatures and support downstream biological interpretation.

Cell-type annotation was performed using a combination of automated and manual approaches. Initial annotation was conducted using SingleR (v2.4.1)^41^, which assigns cell identities based on reference transcriptomic profiles. Automated annotations were subsequently refined through manual inspection of canonical marker genes and cluster specific DEGs. Cell identities were assigned according to established lineage markers, including EPCAM and KRT19 for epithelial cells, COL1A1, COL1A2, DCN, and LUM for fibroblasts, C1QA, C1QB, CD68, and CSF1R for macrophages, CD3D and CD3E for T cells, MS4A1 and CD79A for B cells, CLEC4C and LILRA4 for dendritic cells, PECAM1 and VWF for endothelial cells, TPSAB1 and KIT for mast cells, and PPBP and PF4 for platelets.

To ensure annotation accuracy, automated predictions were cross validated with canonical marker expression patterns visualized by UMAP^42^, feature plots, and differential expression analyses. Clusters exhibiting consistent expression of lineage-specific markers were assigned to the corresponding cell type. This process identified 42 transcriptionally distinct clusters that were consolidated into major cellular compartments, including epithelial cells, fibroblasts, macrophages, dendritic cells, endothelial cells, T cells, B cells, natural killer cells, mast cells, monocytes, neurons, and platelets.

Because stromal and immune remodeling represented the primary focus of this study, fibroblast, macrophage, and dendritic-cell populations were subsequently subjected to detailed differential expression, pathway enrichment, and network analyses to characterize CAF-associated, TAM-associated, and antigen-presentation-related transcriptional programs during PDAC progression.

### 5. Cell Type-Specific KEGG Pathway Enrichment and Network Analysis

To identify the biological processes and signaling pathways that characterize individual cellular populations within the PDAC tumor microenvironment, we performed cell type-specific differential expression analysis followed by Gene Set Enrichment Analysis (GSEA) using Kyoto Encyclopedia of Genes and Genomes (KEGG) pathways. Unlike over-representation analysis, GSEA evaluates the entire ranked gene list and therefore enables detection of coordinated pathway-level changes even when individual genes exhibit modest expression differences.

For each annotated cell type, genes were ranked according to their average log2 fold-change values obtained from differential expression analysis. KEGG pathway enrichment was performed using the gseKEGG function implemented in the clusterProfiler package^43^. Pathways with a false discovery rate (FDR)-adjusted P value < 0.05 were considered significantly enriched. Enrichment scores and normalized enrichment scores (NES) were used to identify the dominant biological programs associated with each cellular population. Enriched KEGG pathways for each cell type are listed in Supplementary Table S3.

Oncogenic pathway activity was evaluated at the single-cell level using four signaling signatures obtained from the Molecular Signatures Database (MSigDB): HALLMARK_KRAS_SIGNALING_UP (KRAS), KEGG_MAPK_SIGNALING_PATHWAY (MAPK), KEGG_MTOR_SIGNALING_PATHWAY (mTOR), and REACTOME_PI3K_AKT_SIGNALING_IN_CANCER (PI3K) (Supplementary Table S4). A score for each signature was calculated for every cell, and scores were compared between each pair of cell types using two-sided Wilcoxon rank-sum tests with adjustment for multiple comparisons (Supplementary Table S3).

To facilitate biological interpretation of stromal and immune remodeling, significantly enriched pathways were further grouped into major functional categories, including extracellular matrix remodeling, cytokine and growth factor signaling, antigen presentation, immune regulation, hypoxia and metabolism, angiogenesis, and cell migration. Particular attention was given to fibroblast, macrophage, dendritic-cell, endothelial-cell, and lymphoid populations because these compartments represented the principal stromal and immune components of the PDAC microenvironment.

To investigate functional interactions among genes associated with stromal and immune remodeling, curated CAF-associated and TAM-associated gene sets were assembled from published literature and matched against the differentially expressed genes identified in the integrated single-cell dataset. Network analyses were used to evaluate the distribution of CAF- and TAM-associated pathways across multiple cellular compartments and to identify signaling programs potentially linking fibroblasts, macrophages, endothelial cells, dendritic cells, and lymphoid populations during PDAC progression.

### 6. Curation of CAF- and TAM-Associated Gene Signatures

To investigate stromal and immune remodeling in PDAC, curated cancer-associated fibroblast (CAF) and tumor-associated macrophage (TAM) gene signatures were assembled from published literature describing fibroblast activation, extracellular matrix remodeling, cytokine signaling, macrophage polarization, immune suppression, angiogenesis, and metabolic adaptation in solid tumors. CAF-associated genes included markers and pathway components related to extracellular matrix organization, collagen remodeling, TGF-β signaling, growth factor signaling, epithelial–mesenchymal transition (EMT), mechanotransduction, hypoxia, and immune regulation. TAM-associated genes included markers associated with macrophage recruitment and survival, CSF1/CSF1R signaling, antigen presentation, phagocytosis, scavenger receptor activity, angiogenesis, hypoxia, glycolytic metabolism, and immunoregulatory macrophage polarization.

The curated CAF and TAM gene sets were subsequently matched against the differentially expressed genes (DEGs) identified from the integrated PDAC single-cell atlas. For each matched gene, cell-type identity, average log2 fold change, expression frequency, and statistical significance were extracted and compiled for downstream analyses. This approach enabled systematic evaluation of the distribution of CAF- and TAM-associated genes across distinct stromal, immune, and endothelial populations. Rather than treating CAF and TAM programs as fibroblast- and macrophage-restricted signatures, this analysis was designed to evaluate the extent to which stromal remodeling pathways were distributed across multiple cellular compartments. The resulting gene signatures and interaction networks were used to characterize multicellular stromal–immune remodeling associated with PDAC progression.

### 7. Clinical specimens

Fresh pancreatic tissue samples from adult patients with intraductal papillary mucinous neoplasms (IPMNs) who underwent surgical resection were collected under Johns Hopkins University Institutional Review Board approval (IRB00282409). The requirement for informed consent was waived for tissue acquisition under this protocol.

The global proteomic cohort included 76 macrodissected normal main pancreatic ducts and 64 IPMNs obtained from patients who underwent pancreatic resection in five countries. Of the 76 normal duct specimens, 32 were obtained from patients with IPMN, whereas 44 were obtained from patients who underwent surgery for pancreatic neoplasms that did not involve the ductal system. Hematoxylin and eosin-stained sections from all IPMNs were reviewed by a pathologist (T.F.) to confirm the diagnosis, direction of differentiation, and histologic grade.

In addition, 104 fresh-frozen PDAC specimens containing more than 15% neoplastic cellularity and 43 matched normal-adjacent tissue (NAT) specimens were obtained from a previously published multi-omics study of 144 PDACs^29,44^. Remaining peptides from these specimens were reanalyzed using the same DIA-MS platform applied to the normal duct and IPMN specimens, in accordance with Clinical Proteomic Tumor Analysis Consortium guidelines. Only global protein-abundance data were included in the present analysis; cyst-fluid, glycoproteomic, genomic, laser-microdissection, and immunolabeling data were not evaluated.

### 8. Global proteomic sample preparation

Fresh-frozen tissue was cryo-pulverized and lysed by three 30-second sonication cycles in ice water in 8 M urea, 75 mM NaCl, 50 mM Tris (pH 8.0), 1 mM EDTA, 2 micrograms/mL aprotinin, 10 micrograms/mL leupeptin, 1 mM PMSF, 10 mM NaF, phosphatase inhibitor cocktails 2 and 3 (each 1:100), and 20 micromolar PUGNAc. Debris was removed at 16,000 x g for 15 minutes at 4 degrees C. Protein concentration was measured by BCA assay and adjusted to 8 mg/mL in urea lysis buffer. Proteins were reduced with 6 mM dithiothreitol for 1 hour at 37 degrees C and alkylated with 12 mM iodoacetamide for 45 minutes at room temperature in the dark. Samples were diluted to 2 M urea with 50 mM Tris-HCl (pH 8.0), digested with Lys-C at 1 mAU per 50 micrograms protein for 2 hours at room temperature, and then digested with trypsin at 1 microgram per 50 micrograms protein overnight at room temperature. Digestion was stopped with 50% formic acid to pH <3. Approximately 5 micrograms of peptide were desalted by stage tip and dried by vacuum centrifugation; 1 microgram was loaded onto an Evotip for global proteomic analysis. No glycopeptide-enrichment step was used for the data analyzed here.

### 9. Liquid chromatography and mass spectrometry

Peptides were separated with an Evosep One system on a 15 cm x 150 micrometer, 1.5 micrometer PepSep C18 column maintained at 50 degrees C using the 30-samples-per-day gradient and analyzed on a timsTOF-HT mass spectrometer. Global bulk proteomic data were acquired in DIA-PASEF mode with an MS1 range of 100-1,700 m/z, an MS2 range of 338-1,338 m/z, 25.0-Da isolation windows without mass overlap, one mobility window, a 1/K0 range of 0.70-1.45 V s/cm2, and an 85-ms ramp time. Samples were randomized and analyzed blinded to diagnosis; repeated pooled samples were included as quality controls throughout data generation.

### 10. Differential expression and stepwise protein selection

To define the full and stepwise directional protein sets analyzed in this report, differential expression between PDAC I/II and non-diseased pancreas (ND) was tested for every protein in the retained proteomic matrix using the two-sided Wilcoxon rank-sum test, with p-values adjusted for multiple testing by the Benjamini-Hochberg method. Of 8,962 proteins tested, 1,072 met the significance criteria of FDR < 0.05 and |log2 fold change| > 1, comprising 586 proteins upregulated and 486 proteins downregulated in PDAC relative to ND. Log2 fold change was calculated as the difference between group medians of log2-transformed abundance, using the samples in which each protein was detected, and results were displayed as a volcano plot of median log2 fold change against −log10(FDR), with the stepwise proteins defined below highlighted (Supplementary Figure S8). This significant set defines the full upregulated and full downregulated feature sets used throughout the report; after excluding proteins with excessive missingness within the five-group comparison (retaining those with no more than 30% missing values, with remaining gaps median-imputed), 487 upregulated and 396 downregulated proteins were carried forward into the full-set PCA and dispersion analyses (Sections 3 and 5).

The stepwise subset was defined by applying a stricter, monotonic-ordering criterion to this same significant protein list, testing whether each protein’s median abundance changed in a single consistent direction across six ordered histopathologic reference groups (ND, normal-adjacent tissue, low-grade IPMN, high-grade IPMN, invasive IPMN, and PDAC I/II): either strictly increasing at every step for upregulated candidates or strictly decreasing at every step for downregulated candidates, with detection in at least 50% of samples in every group. This ordering is an analytical reference structure used to test directional consistency and does not imply that individual patients pass sequentially through each category. Of the 1,072 significant proteins, 68 satisfied this ordering constraint across all six stages-43 upregulated and 25 downregulated-and were retained as the curated stepwise upregulated, stepwise downregulated, and stepwise combined feature sets used in Sections 4 through 6. Because this ordering criterion is more stringent than simple significance, the stepwise sets are, by construction, a subset of the full sets rather than an independently derived group of proteins.

### 11. PCA and Multivariate Dispersion Analysis

Six feature sets were evaluated in this framework: stepwise upregulated proteins (n=43), stepwise downregulated proteins (n=25), stepwise combined proteins (n=68), full upregulated proteins (n=487 retained), full downregulated proteins (n=396 retained), and full combined proteins (n=883 retained), across five diagnostic groups: non-diseased pancreas (ND, n=45), low-grade IPMN (n=30), high-grade IPMN (n=26), invasive IPMN (n=8), and PDAC I/II (n=85). For each feature set, protein abundances were standardized by per-protein z-scoring before principal component analysis, which was used strictly as an unsupervised visualization of global sample structure and not as a classifier; group separation in this PCA-reduced space was then tested by PERMANOVA with 9,999 permutations, assessing whether group centroids differed in multivariate space and yielding R² values describing the proportion of variation associated with diagnostic group, and by PERMDISP, also with 9,999 permutations, testing whether mean distances to group centroids differed among groups-interpreted only as a description of relative within-group heterogeneity, since PERMDISP identifies that dispersion differs without identifying which biological pathway causes it and does not substitute for pairwise dispersion testing. To address the stability of the small invasive-IPMN group (n=8) underlying these dispersion estimates, group-wise bootstrap resampling (2,000 resamples, resampling with replacement within each diagnostic group, followed by re-embedding via principal-coordinates analysis) was used to obtain 95% confidence intervals on each group’s mean centroid distance, and for the six-feature-set summary specifically, dispersion was additionally normalized to the mean dispersion within each feature set, permitting comparison of relative group patterns across feature sets of different size and scale rather than comparing raw centroid distances as though they shared a common absolute scale.

### 12. Identification and evaluation of protein PDAC panel

To identify a reproducible and extracellularly accessible PDAC protein panel, tumor-enriched cell populations were first defined by comparing cell counts between tumor and normal pancreatic tissues using Fisher’s exact test; populations with an odds ratio greater than 1 and a Benjamini–Hochberg-adjusted P value less than 0.05 (natural killer cells, mast cells, fibroblasts, and macrophages) were retained, whereas tumor-depleted populations were excluded. Using the single-cell marker results generated by the workflow described above, all genes that were significant markers of at least one retained population in at least one of four analyses (GSE278694 alone; GSE197177 alone; a three-dataset integration of GSE278694, GSE212966, and GSE217845; and a four-dataset integration additionally including GSE300304) were pooled, yielding 1,927 unique candidates. These candidates were cross-referenced with the global DIA-MS tissue proteomic dataset, in which no glycopeptide-enrichment step was used; 695 candidates had a confidently quantified corresponding protein. For each protein, abundance in PDAC stages I–IV combined (n = 145) was compared separately with non-diseased pancreas (ND, n = 45) and normal-adjacent tissue (NAT, n = 102) using two-sided Mann–Whitney U tests on median-normalized log2 abundance, with Benjamini–Hochberg correction across the proteins tested within each comparison. Proteins were retained only when elevated in PDAC in both comparisons (adjusted P < 0.05 and log2 fold change > 0), leaving 378 candidates. Final selection prioritized concordant marker status across the four single-cell analyses, proteomic detection completeness across all six disease groups, and a documented secreted, cell-surface, or otherwise extracellularly accessible pool based on published literature and UniProt annotation. This procedure identified TIMP1, ANXA2, POSTN, and ISG15: TIMP1, ANXA2, and ISG15 were significant markers in all four single-cell analyses, POSTN was confirmed in three, and all four were quantified in 99.7–100% of proteomic samples. After the panel had been fixed without reference to discriminative performance, each protein was evaluated for discrimination of PDAC from ND and of PDAC from NAT using the same procedure applied in the benchmarking analysis (Section 14). Protein quantities were log2-transformed and median-centered within each sample; the few missing values were set one log2 unit below the lowest observed value of that protein, and AUCs were unchanged under median imputation because all four proteins were detected in at least 99.6% of samples in each comparison. The AUC was computed directly from protein abundance, which for a single protein is equivalent to that of a univariable logistic regression model, and 95% confidence intervals were obtained by stratified bootstrap resampling (2,000 resamples). Sensitivity was estimated in repeated stratified five-fold cross-validation (20 repeats) at a threshold set at the 95th percentile of control scores in each training fold and applied to the held-out fold, and is reported together with the specificity achieved.

### 13. Cellular attribution of stepwise proteins and TAM gene-program analysis

Cell-type markers were taken from the Seurat “FindAllMarkers” analysis of the integrated atlas described above (Supplementary Table S2), which compares each annotated cell type with all remaining cells and reports only genes enriched in that cell type; genes with a Bonferroni-adjusted P value < 0.05 were considered significant markers. Because this analysis identifies enriched but not depleted genes, it cannot assign a cellular source to proteins that decrease in PDAC; cellular attribution was therefore interpreted for the 43 stepwise-increasing proteins. Each of the 68 stepwise proteins was mapped from its UniProt accession to its gene symbol, and the average log2 fold change of that gene in each of the 12 annotated cell types was displayed as a heatmap alongside the protein’s proteomic log2 fold change for PDAC I/II versus ND (Supplementary Figure S9). Stepwise-increasing proteins whose genes were significant fibroblast markers were designated CAF-associated, and those whose genes were significant macrophage markers were designated TAM-associated; proteins meeting both criteria were reported in both groups (Supplementary Figure S10). For the TAM gene-program analysis, TAM-associated genes curated from the literature were grouped into six functional modules (recruitment and survival, immunoregulatory differentiation, hypoxia and metabolic adaptation, inflammatory signaling, CAF–TAM crosstalk, and antigen presentation), and the marker average log2 fold change of each gene in each cell type was displayed; cell types in which a gene was not a significant marker were shown as not enriched (Figure 4).

### 14. Benchmarking against diagnostic IHC markers, molecular-testing-associated proteins, and published IPMN-progression proteins

Candidate proteins and panels were benchmarked in the same DIA-MS dataset against three reference sets: (i) proteins corresponding to diagnostic IHC markers (S100P, IMP3, maspin, MSLN, MUC1, MUC5AC, CK7, CK19, and Ki-67), also combined as S100P+IMP3+maspin+MSLN; (ii) the protein products of KRAS, TP53, SMAD4, and CDKN2A, combined as a single panel, together with Gαs (GNAS); and (iii) the IPMN-progression proteins and combinations reported by Wang et al.²⁹ (TACSTD2, PLOD3, FAM3D, LGALS9, IRS2, PLOD3+FAM3D, LGALS9+IRS2, and LGALS9+TACSTD2), with PLOD3+FN1+TNC+LUM, a combination of invasive-progression proteins from the same study. All combinations were refitted in the present data. Because all markers were analyzed as MS-quantified tissue protein abundance, this comparison does not assess IHC staining, mutation status, or sequencing-based tests. PDAC (n = 145) was compared with low- and high-grade IPMN (n = 56) and with ND (n = 45), excluding invasive IPMN (n = 8).

Preprocessing, single-protein AUCs and confidence intervals, and sensitivity at a cross-validated 95%-specificity threshold followed Section 12, with the direction of each single protein chosen per comparison. Because several reference proteins were detected unevenly between groups (e.g., maspin in 17/45 ND versus 145/145 PDAC samples), each model was also evaluated with median imputation and by complete-case analysis (permitted when at least 10 controls remained), and the highest AUC was reported with its sample size as a best-case benchmark. Models whose AUC varied by more than 0.05 across approaches were flagged, and binary detection scores were used for individual proteins to estimate discrimination attributable to detection alone. Panels were fitted by L2-regularized logistic regression (C = 1; scikit-learn 1.9.1) in repeated stratified five-fold cross-validation (20 repeats), with imputation, scaling, and coefficients estimated within training folds; confidence intervals came from 500 bootstrap resamples with the pipeline refitted and evaluated out-of-bag. AUC differences were tested by paired stratified bootstrap (2,000 resamples) of out-of-fold probabilities for panels or abundance for single proteins, using below-detection imputation on all samples so that compared models shared identical samples.

## Results

### 1. Construction of an Integrated Single-Cell Atlas of Pancreatic Ductal Adenocarcinoma

To characterize cellular heterogeneity within the pancreatic ductal adenocarcinoma (PDAC) tumor microenvironment, we integrated four publicly available treatment-naïve single-cell RNA sequencing datasets (GSE278694, GSE212966, GSE217845, and GSE300304) comprising 26 PDAC patients and 324,662 high-quality cells following quality control, normalization, and batch correction (Figure 1A). Integration using Harmony effectively minimized dataset-specific batch effects while preserving biologically meaningful cellular variation across cohorts.

Unsupervised clustering identified 42 transcriptionally distinct clusters (Figure 1B) that were subsequently consolidated into 12 major cellular populations based on SingleR prediction and canonical marker-gene expression (Figure 1C). These populations included B cells (CD79A, JCHAIN, MS4A1), T cells (CD3D, CD3E, IL7R), natural killer (NK) cells (NKG7, GNLY, GZMB), monocytes (CD14, S100A8, S100A9), macrophages (CD68, C1QC, MARCO), dendritic cells (CLEC4C, LILRA4), fibroblasts (COL1A1, COL1A2, DCN, LUM), epithelial cells (KRT18, KRT19, EPCAM), endothelial cells (CLDN5, PECAM1, VWF), mast cells (TPSAB1, TPSB2, KIT), platelets (PPBP, PF4), and neuronal cells (SOX2, NEFL).

The UMAP embedding demonstrated clear segregation of immune, stromal, epithelial, vascular, and rare cellular compartments, indicating robust transcriptional differences among cell lineages. Immune populations occupied distinct regions of the transcriptional landscape, with T cells, B cells, NK cells, monocytes, macrophages, and dendritic cells forming well-resolved clusters. Stromal populations, including fibroblasts and endothelial cells, exhibited strong separation from immune compartments, while epithelial cells formed a discrete cluster characterized by epithelial lineage markers. Smaller populations of mast cells, platelets, and neuronal cells were also consistently detected, demonstrating the sensitivity of the integration strategy in capturing rare cell populations.

Feature-plot visualization confirmed highly restricted expression of lineage-specific markers within their respective cellular populations (Figure 1D). For example, CD79A expression was confined to B cells, CD3D to T cells, NKG7 to NK cells, COL1A1 to fibroblasts, KRT18 to epithelial cells, and CLEC4C to dendritic cells. Together, these analyses established a comprehensive PDAC single-cell atlas encompassing immune, stromal, epithelial, vascular, and rare cellular compartments, providing the foundation for subsequent analyses of fibroblast activation, macrophage remodeling, dendritic-cell dynamics, and multicellular stromal–immune interactions during PDAC progression.

### 2. Tumor Microenvironment Remodeling During PDAC Progression

To investigate how cellular composition changes during pancreatic ductal adenocarcinoma (PDAC) progression, we first examined the relative abundance of major cell populations across individual patient samples (Figure 2A).

**Figure 2.**
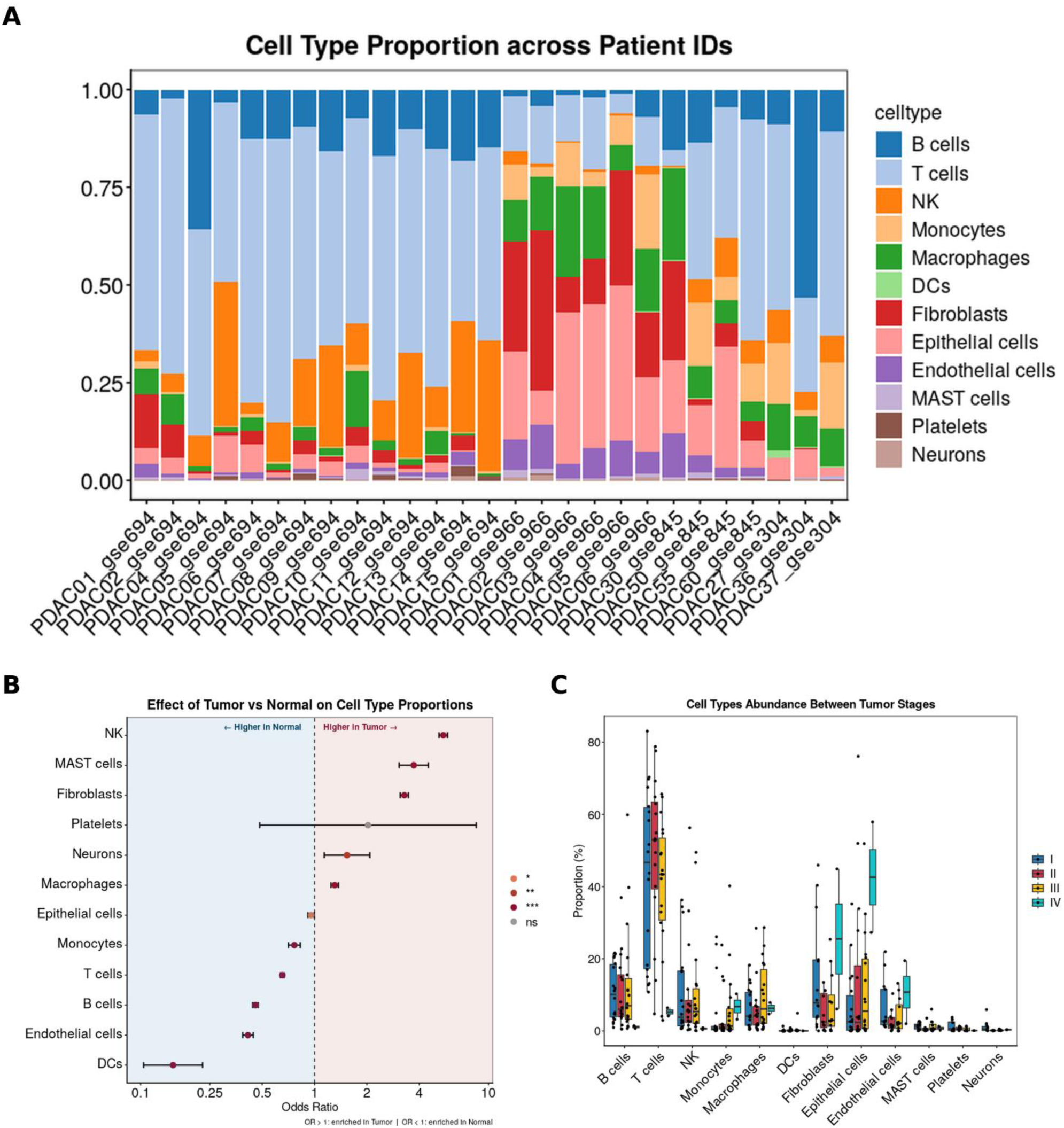
Cell-type composition of PDAC tumors by tissue type and clinical stage. (A) Relative proportion of each major cell type in each patient’s tumor sample (n = 26); the suffix of each sample ID indicates the source dataset. (B) Odds ratios (points) and 95% confidence intervals (bars) for each cell type in tumor versus normal tissue, calculated by Fisher’s exact test on pooled cell counts with Benjamini–Hochberg adjustment; OR > 1 indicates enrichment in tumor (log scale). *P < 0.05, **P < 0.01, ***P < 0.001; ns, not significant. (C) Proportion (%) of each cell type per sample by clinical stage (I–IV). Boxes show the median and interquartile range, whiskers extend to 1.5 × IQR, and points represent individual samples.

Substantial inter-patient heterogeneity was observed, reflecting the diverse cellular architecture of the PDAC tumor microenvironment. While T cells represented the dominant immune population in many samples, several tumors exhibited marked expansion of fibroblasts, epithelial cells, and macrophages. In contrast, dendritic cells were consistently rare across most tumor specimens.

To quantify tumor-associated alterations in cellular composition, we compared tumor and normal tissues using odds ratio analysis (Figure 2B; Supplementary Figure S7). Fibroblasts demonstrated one of the strongest tumor-associated enrichments (OR ≈ 3–4), indicating substantial expansion of stromal populations during tumor development. NK cells and mast cells were also significantly enriched in tumor tissues, whereas macrophages showed moderate enrichment. In contrast, dendritic cells exhibited the most pronounced depletion (OR ≈ 0.15), suggesting a marked reduction in antigen-presenting capacity within the tumor microenvironment. B cells, T cells, and endothelial cells were similarly reduced in tumors relative to normal pancreatic tissue. Monocytes were also modestly reduced in tumors, neurons were modestly enriched, and epithelial cells showed a small but significant reduction, whereas platelet abundance did not differ significantly between tumor and normal tissue.

Next, we examined changes in cellular abundance across clinical stages (Figure 2C). Because the cohort included only one stage IV patient, these stage-level comparisons are descriptive and were not tested statistically. Fibroblast and epithelial-cell proportions were highest in the stage IV sample, whereas stages I–III showed broadly overlapping distributions, and the median fibroblast proportion in stage III was not higher than in stage I. Macrophage proportions were highest in stage III, and monocyte and endothelial-cell proportions were higher in stage IV than in earlier stages. In contrast, T-cell, B-cell, and NK-cell proportions were lower in the stage IV sample than in stages I–III, while dendritic cells, mast cells, platelets, and neurons remained rare at every stage. Together, these observations suggest that advanced disease may be accompanied by expansion of stromal and epithelial compartments at the expense of lymphoid populations, a pattern that requires confirmation in cohorts with more advanced-stage samples.

To determine whether these compositional changes were accompanied by distinct biological programs, we performed cell-type-specific KEGG pathway enrichment analysis (Figure 3A). Fibroblasts were enriched for focal adhesion, ECM–receptor interaction, PI3K–Akt signaling, integrin signaling, cytoskeleton in muscle cells, and protein digestion and absorption, consistent with activation of cancer-associated fibroblast (CAF) programs. Each of these programs has an established link to PDAC biology: focal adhesion kinase activity sustains the fibro-inflammatory PDAC stroma, and its inhibition reduces fibrosis and sensitizes tumors to checkpoint immunotherapy^45^; beta-1 integrin mechanosignaling and matricellular fibrosis set tumor tissue tension and are associated with shorter patient survival^46^; and the PI3K p110-alpha isoform is required for oncogenic KRAS-driven acinar-to-ductal metaplasia and PDAC formation^47^. T cells were enriched for T-cell receptor signaling, PD-L1 expression and PD-1 checkpoint signaling, Th17 and Th1/Th2 cell differentiation, and oxidative phosphorylation. The PD-1/PD-L1 axis is of particular interest in this disease, where single-agent and dual checkpoint blockade have produced minimal objective responses^11,48^, and pathogenic Th17 rather than Th1 immunity has been linked to PDAC progression^49^. B cells were enriched for antigen processing and presentation, phagosome, hematopoietic cell lineage, and the intestinal immune network for IgA production. Consistent with a functional role for B cells in this setting, the spatial distribution of tumor-infiltrating B cells predicts prognosis in human pancreatic adenocarcinoma^50^. Macrophages were enriched for tight junction and a group of immune-mediated disease terms (systemic lupus erythematosus, allograft rejection, graft-versus-host disease, autoimmune thyroid disease, and asthma); because these KEGG terms share MHC class II genes, this pattern is consistent with an antigen-presenting macrophage state. MHC class II-expressing myeloid populations are a prominent feature of the PDAC leukocyte landscape and have been associated with patient outcome^32^. Terms enriched in the small neuronal population were dominated by neurodegeneration pathways (Parkinson, Huntington, Alzheimer, and prion disease, and amyotrophic lateral sclerosis) and oxidative phosphorylation, which share mitochondrial respiratory-chain genes. Mitochondrial respiration is itself a relevant axis in PDAC, because tumor cells that survive KRAS pathway inhibition depend on oxidative phosphorylation and are sensitive to its inhibition^51^, and neural signaling is increasingly recognized as an active component of the PDAC microenvironment^15,52^. Dendritic cells showed enrichment of oxidative phosphorylation, whereas the only term detected for NK cells (natural killer cell-mediated cytotoxicity) did not reach the FDR < 0.05 threshold. Integration of cell-abundance and pathway analyses identified three cellular compartments that exhibited the most prominent remodeling in PDAC: cancer-associated fibroblasts (CAFs), tumor-associated macrophages (TAMs), and dendritic cells (DCs). CAFs represented one of the most strongly enriched populations within tumors (OR ≈ 3.2) and showed enrichment of ECM-remodeling, focal adhesion, integrin, and PI3K–Akt signaling pathways. Macrophages were significantly expanded (OR ≈ 1.3) and exhibited a transcriptional program characterized by C1QA, C1QB, CD163, MSR1, FCGR3A, and LILRB4 expression, consistent with immunoregulatory TAM differentiation. A complement-high, C1QA+ macrophage state with these features has been reported repeatedly in PDAC and in pan-cancer myeloid atlases^25,30^. In contrast, dendritic cells showed the strongest depletion among all cell populations (OR ≈ 0.15), suggesting impaired antigen-presenting capacity and diminished antitumor immune surveillance. Dendritic cell paucity has been shown experimentally to produce dysfunctional immune surveillance and to favor tumor-promoting Th17 responses in PDAC, and restoring conventional dendritic cells restrains disease^49^. Although NK cells exhibited the highest degree of tumor enrichment overall (OR ≈ 5–6), NK-cell proportions were lower in the stage IV sample than at earlier stages, and the only KEGG term detected for NK cells did not reach significance; NK-cell biology was therefore not pursued further in the present study. NK cells in PDAC are additionally suppressed at the functional level by tumor-derived extracellular vesicles, so abundance alone may not reflect cytotoxic capacity^53^.

**Figure 3.**
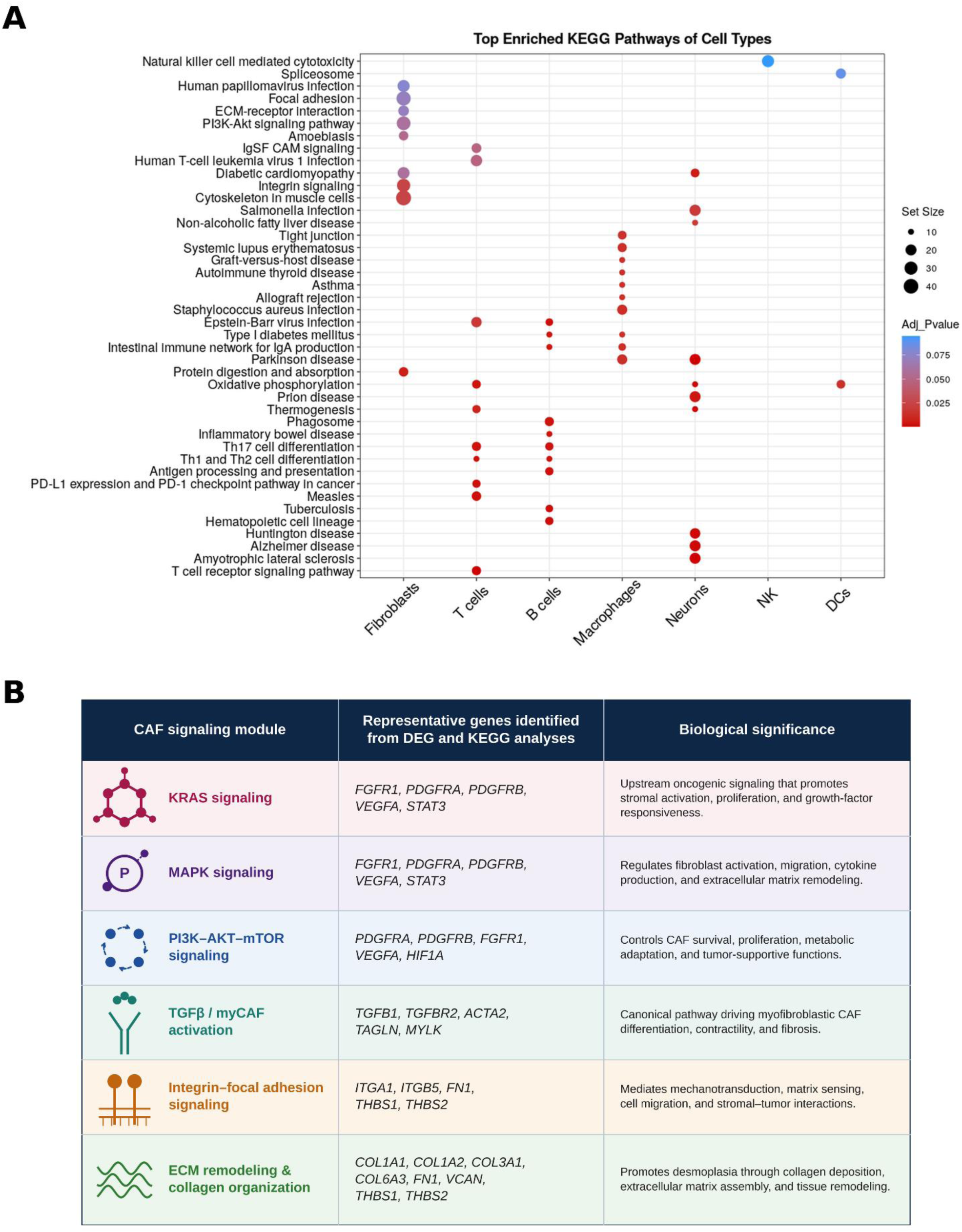
Cell-type pathway enrichment and fibroblast signaling programs in the PDAC microenvironment. (A) Top enriched KEGG pathways for each cell type from gene set enrichment analysis (gseKEGG) of genes ranked by average log2 fold change relative to all other cells. Dot size indicates set size (number of pathway genes in the ranked list), and color indicates the Benjamini–Hochberg-adjusted P value. (B) Literature-curated CAF signaling modules, representative genes identified in the fibroblast differential expression and KEGG analyses, and their proposed biological significance.

Together, these observations indicate that PDAC progression is associated with coordinated expansion of stromal and immunosuppressive myeloid compartments accompanied by depletion of antigen-presenting immune populations. The concurrent enrichment of fibroblasts and macrophages, together with the marked depletion of dendritic cells, suggested that PDAC progression is driven by coordinated stromal–immune remodeling rather than isolated changes in individual cell populations. Therefore, subsequent analyses focused on CAFs, TAMs, and DCs to investigate the molecular mechanisms linking stromal activation, macrophage polarization, impaired antigen presentation, and tumor progression within the PDAC microenvironment.

### 3. CAF-associated signaling programs are distributed across multiple cell populations in the PDAC microenvironment

Fibroblast-specific differential expression analysis followed by KEGG enrichment identified significant enrichment of ECM–receptor interaction, focal adhesion, integrin signaling, PI3K–Akt signaling, protein digestion and absorption, and cytoskeletal remodeling pathways (Figure 3A). Collectively, these pathways support a core desmoplastic CAF program characterized by extracellular matrix remodeling, matrix adhesion, mechanotransduction, and PI3K–Akt-mediated survival signaling. In addition to the directly enriched KEGG pathways, several CAF-relevant signaling modules, including KRAS signaling, MAPK signaling, and TGFβ/myCAF activation, were supported by expression of canonical pathway-associated genes identified in the fibroblast DEG analysis, including FGFR1, PDGFRA, PDGFRB, VEGFA, STAT3, TGFB1, TGFBR2, ACTA2, and TAGLN. These pathways were therefore retained as biologically relevant CAF signaling axes despite not emerging among the top enriched KEGG terms. Differential expression analysis identified a robust fibroblast-specific transcriptional program characterized by marked enrichment of extracellular matrix (ECM), stromal remodeling, and signaling-associated genes (Figure 3B). In addition to classical CAF markers including ASPN, MFAP4, PRRX1, DCN, SFRP2, PRELP, LUM, COL1A2, COL3A1, BGN, and TAGLN, pathway-focused analysis identified enrichment of genes involved in ECM remodeling (COL1A1, COL1A2, THBS2, VCAN, ACTA2, FAP, and MMP2), growth factor signaling (PDGFRA, PDGFRB, FGFR1, and TGFB1), mechanotransduction (WWTR1, CCN1, and CCN2), epithelial–mesenchymal transition (ZEB1 and ZEB2), hypoxia and metabolic adaptation (HIF1A, PKM, and LDHA), and inflammatory signaling pathways including STAT3, NFKB1, SOCS3, and CCL5.

To investigate the biological programs underlying fibroblast activation and stromal remodeling, we curated CAF-associated signaling pathways from the literature and mapped the corresponding genes onto the integrated single-cell PDAC atlas. Heatmap visualization revealed that classical extracellular matrix (ECM) and myofibroblastic CAF genes, including **COL1A1, COL1A2, FN1, THBS2, POSTN, CTHRC1, ACTA2, FAP, and PDGFRA**, were predominantly enriched within fibroblasts, consistent with activation of a desmoplastic CAF phenotype (Supplementary Table S2).

However, several CAF-associated signaling genes were not restricted to fibroblasts and were detected across multiple stromal and immune cell populations. Components of **TGF-β signaling** (TGFB1, TGFBR2), **KRAS/MAPK signaling** (FGFR1, VEGFA, STAT3), **PI3K–AKT–mTOR/hypoxia signaling** (HIF1A, PIK3R1, PKM), and **EMT-associated transcriptional programs** (ZEB1, ZEB2, VIM) were observed in endothelial cells, macrophages, monocytes, and additional immune populations. This distribution is consistent with established PDAC biology: TGF-beta signaling drives the myofibroblastic CAF state and is antagonized by IL-1-induced JAK/STAT signaling, which sustains inflammatory CAFs^54,55^, with TGF-beta-driven LRRC15+ myofibroblasts emerging as a determinant of response to immunotherapy^56^; STAT3 activity in the PDAC epithelium and stroma controls matricellular fibrosis and tissue tension^46^; HIF1A expression in tumor-associated macrophages suppresses T-cell function and promotes tumor progression^28^; and ZEB1 is a key determinant of cellular plasticity and metastasis in this disease ^57^. Similarly, antigen-presentation genes (**CD74, HLA-DRA, HLA-DRB1**) and inflammatory regulators (**CXCL16, IL6ST, SOCS3, NFKB1, HMOX1**) were primarily enriched within myeloid populations but overlapped with CAF-associated signaling networks. Overlapping expression of MHC class II genes between myeloid cells and fibroblasts has been described previously in PDAC, where antigen-presenting CAFs express CD74 and HLA class II genes^12,58^.

Consistent with these observations, pathway activity analysis demonstrated significant enrichment of major CAF-associated oncogenic pathways across multiple cell types (Supplementary Table S3). **KRAS signaling** activity was highest in macrophages and endothelial cells, while **MAPK signaling** was broadly activated across stromal and immune compartments. **PI3K signaling** and **mTOR signaling** were similarly distributed among fibroblasts, macrophages, endothelial cells, and lymphoid populations. Together, these findings indicate that canonical CAF pathways are not confined to fibroblasts but instead operate within a multicellular stromal ecosystem involving endothelial, myeloid, and immune cells. A comparable conclusion has been drawn from spatially confined sub-tumor microenvironments in PDAC, in which stromal and immune programs are organized into shared niches rather than segregated strictly by cell type^26^.

These results support a model in which CAF-associated signaling represents a coordinated tissue-level remodeling program rather than an isolated fibroblast phenotype. ECM-producing fibroblasts likely provide the structural foundation of the desmoplastic microenvironment, whereas macrophages, endothelial cells, and immune populations contribute complementary signaling activities through KRAS/MAPK, PI3K–mTOR, hypoxia, inflammatory, and antigen-presentation pathways. The convergence of these pathways across multiple cell populations indicates that these signaling activities are distributed across multiple compartments rather than restricted to a single cell type, consistent with broad stromal-immune remodeling that may collectively contribute to immune suppression, tissue remodeling, and tumor progression in PDAC. Such distributed signaling is itself an obstacle to compartment-directed therapy, because inhibiting a pathway in one cell type may be compensated by the others that express it^14,59,60^

#### Expansion of C1QA⁺ Immunoregulatory Tumor-Associated Macrophages During PDAC Progression

To further characterize the macrophage compartment, TAM-associated genes were curated from the literature and organized into functional pathway modules representing macrophage recruitment and survival, immunoregulatory differentiation, hypoxia and metabolic adaptation, inflammatory signaling, CAF–TAM crosstalk, and antigen presentation. Heatmap visualization of TAM-associated genes across all major cell populations revealed that several hallmark TAM genes were highly enriched within macrophages, whereas additional signaling mediators were preferentially expressed by other cell types (Figure 4).

**Figure 4.**
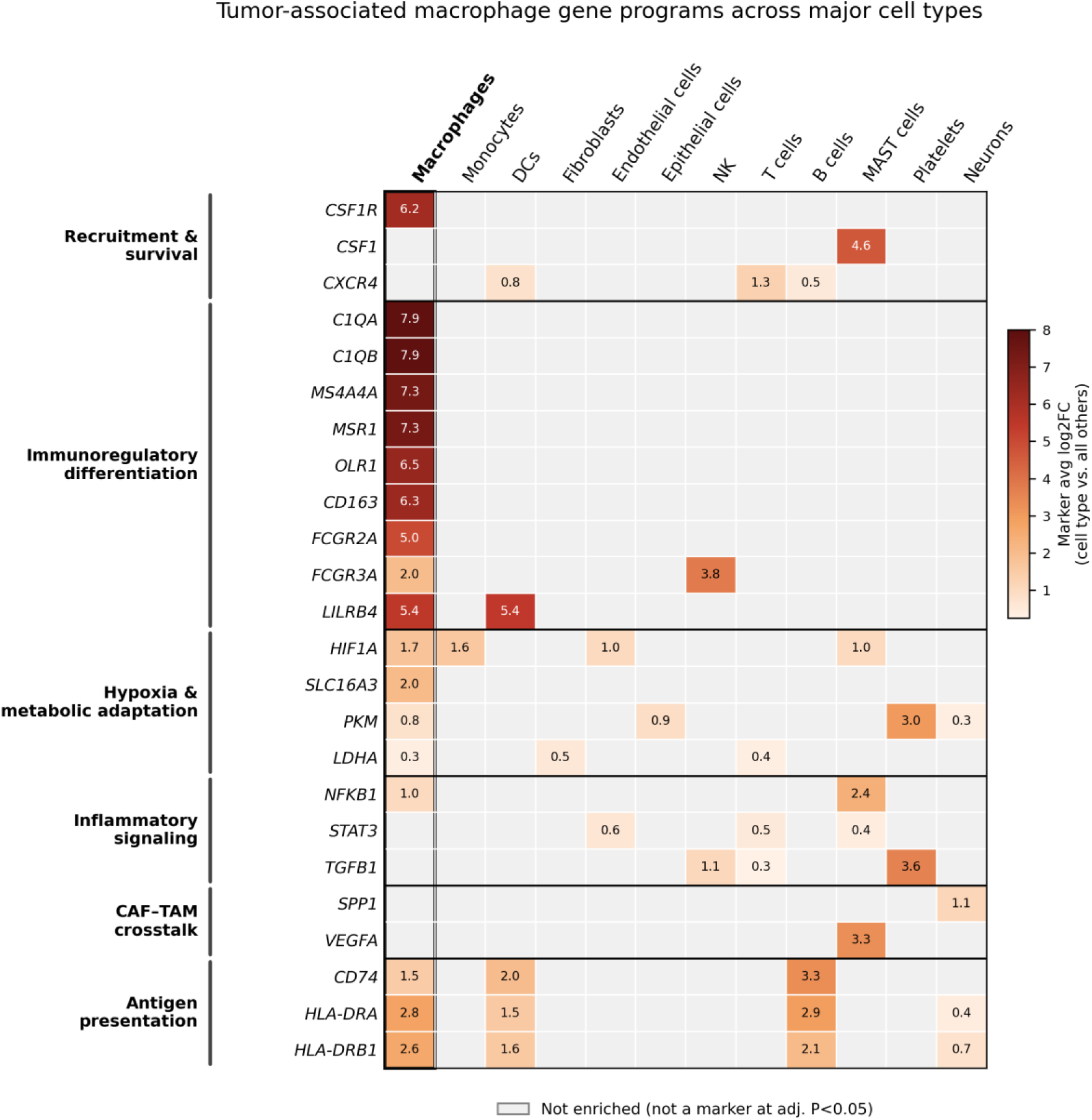
Tumor-associated macrophage gene programs across major cell types. Heatmap of curated TAM-associated genes grouped by functional module (recruitment and survival, immunoregulatory differentiation, hypoxia and metabolic adaptation, inflammatory signaling, CAF–TAM crosstalk, and antigen presentation), Values and color indicate the average log2 fold change of each gene in the indicated cell type relative to all other cells (Wilcoxon rank-sum test, Bonferroni-adjusted P < 0.05); grey indicates that the gene was not a significant marker of that cell type.

Differential expression analysis demonstrated that macrophages possessed one of the most distinct transcriptional programs within the integrated PDAC atlas. The macrophage compartment was dominated by a C1QA⁺/C1QB⁺ immunoregulatory signature, with C1QA (log2FC = 7.86) and C1QB (log2FC = 7.86) representing the most strongly enriched macrophage markers. Additional highly expressed TAM-associated genes included MS4A4A (7.31), MSR1 (7.30), OLR1 (6.47), CD163 (6.31), CSF1R (6.17), and LILRB4 (5.44), supporting the presence of a specialized tumor-associated macrophage population characterized by scavenger receptor activity, complement signaling, and immune regulation.

Module-level analysis showed that macrophage enrichment differed across TAM programs (Figure 4). In the recruitment and survival module, CSF1R was restricted to macrophages (log2FC = 6.17), whereas its ligand CSF1 was enriched in mast cells (4.64) and CXCR4 in T cells, B cells, and dendritic cells, suggesting paracrine support of macrophage recruitment by other immune populations. The immunoregulatory differentiation module was the most macrophage-specific: C1QA, C1QB, MS4A4A, MSR1, OLR1, CD163, and FCGR2A were enriched exclusively in macrophages (log2FC 5.00–7.86), whereas LILRB4 was equally enriched in macrophages and dendritic cells (5.44 and 5.42), and FCGR3A was more strongly enriched in NK cells (3.77) than in macrophages (2.00). Hypoxia and metabolic genes showed only modest macrophage enrichment; SLC16A3 (2.04) was macrophage-restricted, whereas HIF1A (1.73), PKM (0.76), and LDHA (0.34) were also enriched in other cell types, with PKM most enriched in platelets (3.02). Among inflammatory mediators, NFKB1 was enriched in macrophages (0.98) but more strongly in mast cells (2.40), STAT3 was enriched in endothelial cells, T cells, and mast cells, and TGFB1 was enriched in platelets, NK cells, and T cells rather than in macrophages.

The CAF–TAM crosstalk mediators were not enriched in macrophages: SPP1 was a significant marker only of neurons (1.14) and VEGFA only of mast cells (3.28). Antigen-presentation genes CD74, HLA-DRA, and HLA-DRB1 were enriched in macrophages (1.48–2.78), dendritic cells (1.55–1.96), and B cells (2.06–3.32), consistent with a shared professional antigen-presentation program. Because marker testing identifies only cell-type-enriched genes, these results indicate where each gene is preferentially expressed but do not exclude lower-level expression in other compartments.

Collectively, these findings identify a dominant C1QA⁺ immunoregulatory TAM population defined by complement, scavenger-receptor, and Fc-receptor genes that were largely restricted to macrophages. In contrast, several ligands and signaling mediators associated with TAM function, including CSF1, CXCR4, STAT3, TGFB1, SPP1, and VEGFA, were preferentially expressed by other immune or stromal populations rather than by macrophages themselves. Similar to the CAF-associated signaling programs described in Figure 3, these observations suggest that TAM-associated programs in PDAC depend on signals supplied by multiple cell types. The interpretation of this division, and its implications for macrophage-directed therapy, are developed in the Discussion.

### 3. Depletion of Plasmacytoid Dendritic Cells Reveals a Potential Immune-Evasion Mechanism in PDAC

Among all immune populations identified in the integrated PDAC single-cell RNA sequencing dataset, dendritic cells exhibited the most pronounced depletion within tumor tissues compared with adjacent normal pancreas. Quantitative cell abundance analysis demonstrated a marked reduction in dendritic cell representation in tumors, indicating substantial disruption of antigen-presenting cell populations during PDAC progression. Notably, dendritic cell depletion was more prominent than the reduction observed for B cells, T cells, or endothelial cells, suggesting that loss of dendritic-cell-mediated immune surveillance may represent a central feature of PDAC immune remodeling.

| Gene | avg_log2FC | pct.1 | pct.2 | p_val_adj | cluster |
| --- | --- | --- | --- | --- | --- |
| CLEC4C | 9.797157 | 0.718 | 0.001 | 0 | DCs |
| LILRA4 | 8.679508 | 0.858 | 0.006 | 0 | DCs |
| PLD4 | 6.545539 | 0.895 | 0.036 | 0 | DCs |

Transcriptional analysis revealed that the dendritic cell compartment was dominated by a highly specific plasmacytoid dendritic cell (pDC) signature. The most significantly enriched genes included CLEC4C (log2FC = 9.80), LILRA4 (log2FC = 8.68), and PLD4 (log2FC = 6.55), all of which are established markers of human pDCs. These genes were expressed in most dendritic cells but showed minimal expression in other cellular populations, confirming the identity of this cluster as plasmacytoid dendritic cells. Specifically, CLEC4C was detected in 71.8% of dendritic cells compared with only 0.1% of other cells, while LILRA4 and PLD4 were expressed in 85.8% and 89.5% of dendritic cells, respectively, with negligible expression outside the pDC compartment.

The combination of strong lineage-specific marker expression and substantial reduction in overall abundance suggests that pDCs are not merely transcriptionally altered but are depleted in PDAC tumors. This observation identifies loss of pDCs as a distinct component of PDAC-associated immune remodeling. Because pDCs are major producers of type I interferons and play essential roles in antigen presentation and T-cell priming, their depletion may contribute to the defective antitumor immune responses observed in PDAC. Together, these findings identify pDC loss as a potentially underappreciated component of PDAC immune remodeling.

Dendritic cells play an essential role in orchestrating antitumor immunity through antigen presentation, activation of adaptive immune responses, and priming of cytotoxic T lymphocytes. Previous studies have reported global dendritic cell deficiency in PDAC; however, our analysis further refines this observation by demonstrating that the depleted dendritic cell population is predominantly composed of plasmacytoid dendritic cells characterized by expression of CLEC4C, LILRA4, and PLD4.

Plasmacytoid dendritic cells are specialized immune cells responsible for rapid production of type I interferons in response to cellular stress and pathogenic signals. Beyond their role in innate immunity, pDCs contribute to antigen presentation, crosstalk with conventional dendritic cells, and activation of T-cell-mediated immune responses. Therefore, depletion of pDCs may have profound consequences for immune surveillance in PDAC. Loss of pDC-derived interferon signaling may impair recruitment and activation of effector immune cells while simultaneously weakening antigen presentation and T-cell priming within the tumor microenvironment.

Importantly, pDC depletion occurred in parallel with expansion of two dominant immunosuppressive populations identified in our dataset: activated cancer-associated fibroblasts and C1QA⁺ tumor-associated macrophages. The fibroblast compartment exhibited strong enrichment of desmoplastic and extracellular matrix-associated genes, including ASPN, DCN, LUM, COL1A2, and COL3A1, consistent with extensive stromal remodeling. Concurrently, macrophages displayed a highly enriched C1QA⁺/C1QB⁺/CD163⁺/CSF1R⁺ transcriptional program associated with immunoregulatory tumor-associated macrophages. These macrophages additionally expressed the scavenger receptors MSR1 and OLR1 as well as the inhibitory myeloid checkpoint receptor LILRB4, indicating a phenotype specialized for immune suppression, lipid handling, and tumor support.

Activated ASPN⁺/COL1A2⁺/LUM⁺ cancer-associated fibroblasts, C1QA⁺/CD163⁺/CSF1R⁺ tumor-associated macrophages, and depletion of CLEC4C⁺/LILRA4⁺ plasmacytoid dendritic cells emerged concurrently across the integrated PDAC single-cell atlas, suggesting coordinated stromal and immune remodeling rather than independent cellular alterations. Fibroblasts exhibited exceptionally strong activation of extracellular matrix and desmoplastic programs, characterized by marked enrichment of ASPN, DCN, LUM, COL1A2, COL3A1, and BGN, whereas macrophages were dominated by an immunoregulatory C1QA⁺/C1QB⁺ transcriptional program associated with CD163, CSF1R, MSR1, OLR1, and LILRB4 expression. In parallel, dendritic cells represented the most depleted immune population and were characterized by a highly specific plasmacytoid dendritic cell signature defined by CLEC4C, LILRA4, and PLD4. Together, these observations suggest that desmoplastic CAF activation, macrophage enrichment, and dendritic-cell depletion occur concurrently during PDAC progression and may collectively contribute to an immunosuppressive tumor microenvironment. The combined effects of stromal remodeling, expansion of immunosuppressive macrophages, and loss of pDC-associated immune surveillance are predicted to reduce effective T-cell priming and facilitate immune escape during PDAC progression. Taken together, these findings support a model in which PDAC progression is characterized by coordinated stromal and immune remodeling rather than isolated cellular changes. Activated fibroblasts may establish a dense desmoplastic barrier that limits immune-cell infiltration, while C1QA⁺ macrophages create an immunosuppressive environment through checkpoint signaling and metabolic adaptation. Simultaneously, depletion of plasmacytoid dendritic cells may reduce antigen presentation capacity and impair type I interferon-mediated immune activation. The convergence of these processes is predicted to suppress effective antitumor immunity and facilitate immune escape.

### 6. Integrated pathway analyses characterize stromal and immune remodeling in PDAC

To investigate the biological mechanisms underlying the coordinated changes observed across fibroblasts, macrophages, and dendritic cells, we performed pathway enrichment analysis and oncogenic pathway signature scoring across major cellular populations (Figure 3A; Supplementary Table S3).

KEGG pathway enrichment analysis demonstrated functional specialization among major cell populations (Figure 3A). Fibroblasts were enriched for ECM–receptor interaction, focal adhesion, integrin signaling, cytoskeletal, and PI3K–Akt pathways, consistent with the activated desmoplastic CAF phenotype described above. Macrophages were enriched for tight junction and MHC class II-associated immune-disease terms, consistent with an antigen-presenting state, and T cells were enriched for T-cell receptor signaling, PD-L1/PD-1 checkpoint signaling, Th1/Th2/Th17 differentiation, and oxidative phosphorylation, indicating adaptive immune activation accompanied by checkpoint engagement. The dendritic-cell compartment, although depleted in tumors, showed enrichment of oxidative phosphorylation; no antigen-presentation term reached significance in this small population.

To determine whether stromal and immune populations actively participate in tumor-associated signaling programs, KRAS, MAPK, mTOR, and PI3K pathway signature scores were calculated at the single-cell level (Supplementary Table S3). Macrophages exhibited among the highest KRAS, MAPK, and PI3K signaling activities across all cell populations, whereas fibroblasts demonstrated elevated PI3K and mTOR pathway activity. These findings indicate that both CAFs and TAMs actively engage oncogenic signaling pathways commonly associated with PDAC progression rather than functioning as passive components of the tumor microenvironment. Integrated pathway analyses further characterized the functional programs associated with stromal and immune remodeling in PDAC. Fibroblasts were enriched for extracellular-matrix organization, focal adhesion, integrin signaling, and PI3K–Akt pathways, consistent with their desmoplastic phenotype. Macrophages showed an antigen-presentation-associated pathway profile together with increased KRAS, MAPK, and PI3K pathway activity, consistent with the immunoregulatory TAM phenotype identified in the integrated atlas. Dendritic cells, although depleted in tumors, retained a highly specific pDC transcriptional identity (Figure 5).

**Figure 5.**
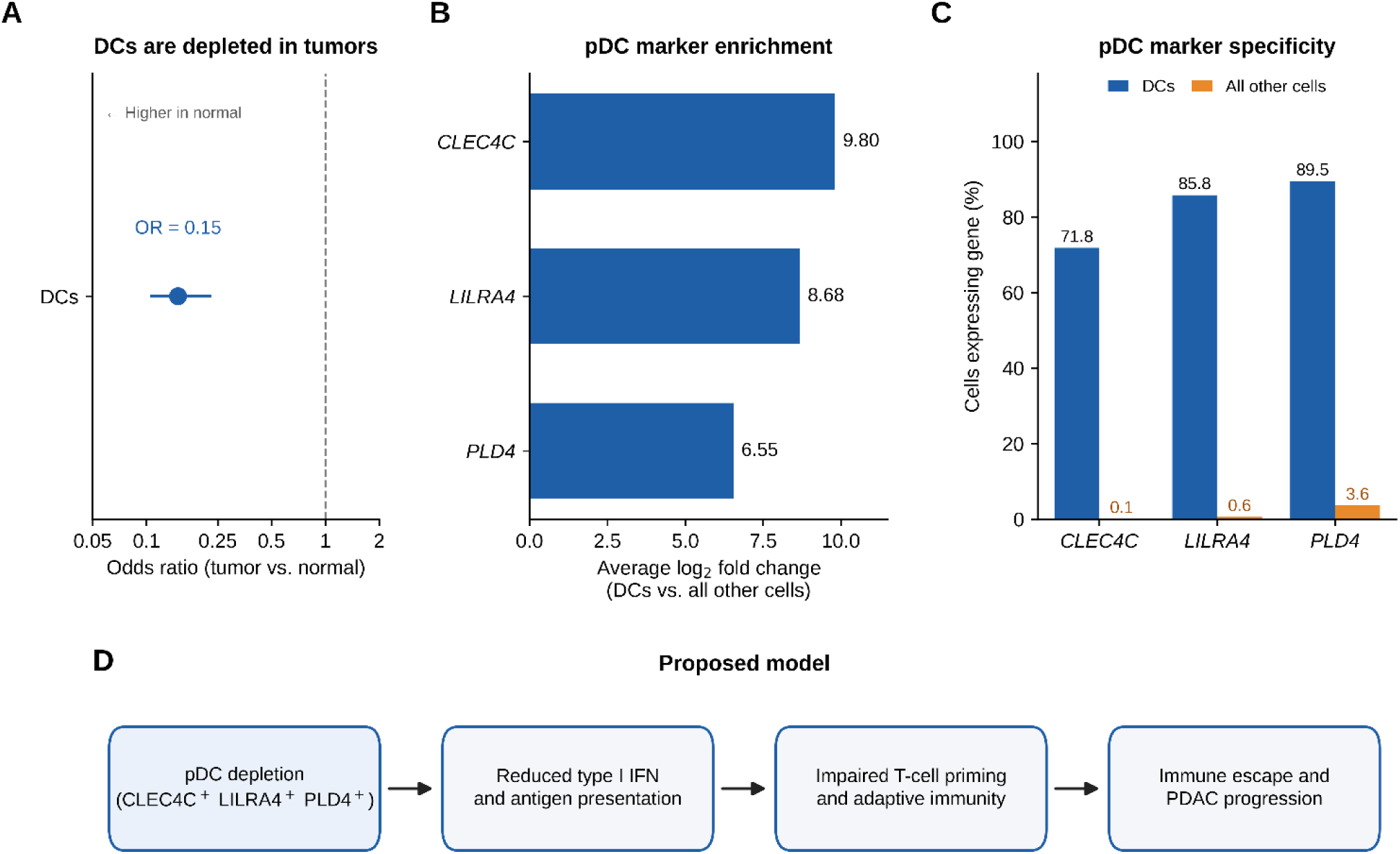
Plasmacytoid dendritic cell signature of the depleted DC compartment in PDAC. (A) Odds ratio (95% CI) for DC abundance in tumor versus normal tissue (Fisher’s exact test). (B) Average log₂ fold change of pDC markers in DCs relative to all other cells. (C) Percentage of DCs versus all other cells expressing each marker. (D) Proposed model linking pDC loss to impaired antitumor immunity. Panel D is a hypothesized model that was not directly tested in this study.

Taken together, these findings demonstrate that PDAC is characterized by concurrent remodeling of stromal and immune compartments involving CAF activation, C1QA+ macrophage enrichment, and pDC depletion. These independently defined cellular programs provide the biological context for the subsequent proteomic analyses and candidate biomarker selection.

#### Stepwise proteomic candidates and their cellular sources

Differential abundance analysis of the tissue proteome identified 1,072 proteins that differed between PDAC I/II (n = 85) and non-diseased pancreas (n = 45) (FDR < 0.05, |log2FC| > 1), of which 586 were increased and 486 decreased in PDAC (Supplementary Figure S8). Of these, 68 (6.3%) also followed the stepwise pattern across the six ordered reference groups, 43 increasing and 25 decreasing. Stepwise-increasing proteins were dominated by extracellular-matrix, protease, and interferon-associated proteins, including THBS2 (log2FC = 5.31), SFRP2 (2.97), LAMC2 (2.47), TYMP (2.46), TIMP1 (2.40), cathepsin S (CTSS; 2.39), and ISG15 (1.81), whereas stepwise-decreasing proteins were dominated by acinar digestive enzymes and mitochondrial metabolic proteins, including AMY2A (−6.73), CELA2A (−4.86), LMF1 (−2.20), and METTL7A (−1.72).

To relate the stepwise proteins to the cellular programs identified in the single-cell atlas, each protein was matched to cell-type markers (Supplementary Figure S9). Twenty-six of the 68 stepwise proteins were significant markers of at least one annotated cell type, and 20 were markers of fibroblasts or macrophages, 18 of which were in the stepwise-increasing set. Only three of the 25 stepwise-decreasing proteins (HERPUD1, NDUFC2, and SEC11C) were markers of any cell type, consistent with the loss of acinar and metabolic function that the decreasing set largely reflects and with the absence of a separately annotated acinar population in the atlas. Forty-two stepwise proteins, including the panel constituents FAT1 and METTL7A, were not markers of any annotated cell type, so their cellular source cannot be inferred from these data.

Among the 43 stepwise-increasing proteins, 9 were fibroblast (CAF) markers and 12 were macrophage (TAM) markers, with three proteins shared between the two sets (TIMP1, ISG15, and CTSB; Supplementary Figure S10). CAF-associated proteins were highly fibroblast-specific: SFRP2 (scRNA-seq log2FC = 8.22) and THBS2 (7.64) were enriched exclusively in fibroblasts, IGFBP5 (6.96) was otherwise enriched only in neurons, and THBS2 was also the most strongly elevated stepwise protein in tissue (proteomic log2FC = 5.31). TIMP1 was predominantly fibroblast-associated (4.08 versus 1.95 in macrophages). By contrast, TAM-associated proteins were shared with other myeloid populations: CTSB (3.98) and TYMP (3.11) were also enriched in monocytes and dendritic cells, CTSS was more strongly enriched in monocytes (3.80) than in macrophages (2.46), and YWHAH and ARPC1B were most strongly enriched in platelets, which in this atlas derive almost entirely from PBMC samples (Supplementary Figure S7). ISG15 was equally enriched in fibroblasts and macrophages (1.06 and 1.03). Thus, the stepwise proteomic signal is linked most specifically to the CAF compartment, whereas TAM-linked proteins reflect a broader myeloid program.

### 7. Identification of protein panels for PDAC detection

The four candidate panels were evaluated by cross-validated logistic regression in two comparisons, PDAC versus low- and high-grade IPMN and PDAC versus non-diseased pancreas (ND), using the procedure described in Section 14 (Figure 6; Table 1). All panels discriminated PDAC from ND almost perfectly (AUC 0.98–1.00), whereas discrimination from precursor lesions separated them more clearly. CATB+EPIPL performed best against IPMN (AUC 0.98, 95% CI 0.95–1.00; sensitivity 92% at 92% specificity) and was the only panel to exceed the refitted PLOD3+FN1+TNC+LUM combination of IPMN-progression proteins (AUC 0.91, 95% CI 0.84–0.99; ΔAUC 0.06, P = 0.004). METTL7A+ISG15 reached an AUC of 0.94 (95% CI 0.89–1.00) with a sensitivity of 71% at 92% specificity, and FAT1+SEC11C reached 0.91 (95% CI 0.85–0.97); neither differed significantly from PLOD3+FN1+TNC+LUM (P = 0.32 and P = 0.94, respectively).

**Figure 6.**
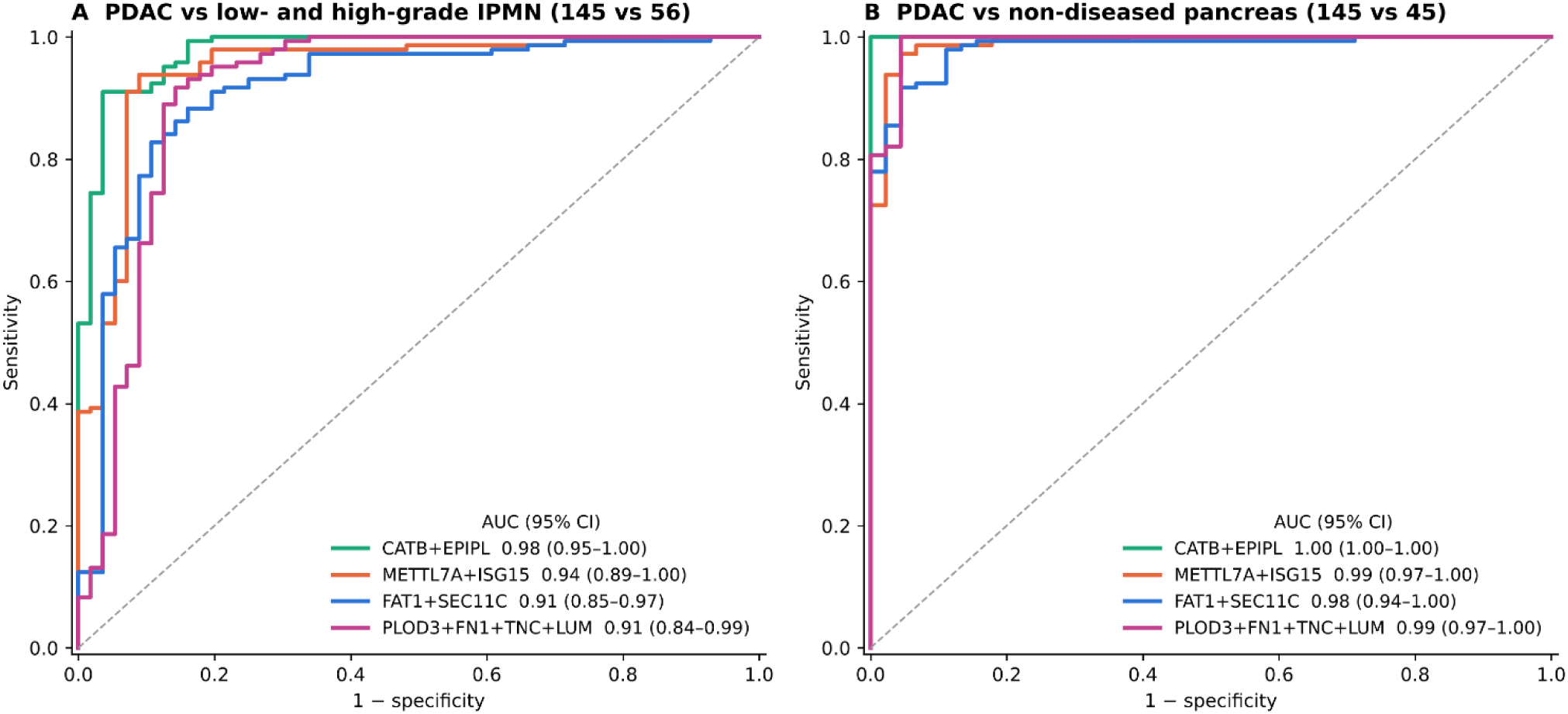
ROC curves of candidate protein panels. Discrimination of PDAC (n = 145) from (A) low- and high-grade IPMN (n = 56) and (B) non-diseased pancreas (n = 45) by CATB+EPIPL, METTL7A+ISG15, FAT1+SEC11C, and PLOD3+FN1+TNC+LUM. Curves are based on out-of-fold predicted probabilities from L2-regularized logistic regression in repeated five-fold cross-validation (×20); AUC (95% CI from out-of-bag bootstrap with model refitting) is shown in the key.

| Panel | Comparison | n (group 1 vs group 2) | AUC (95% CI) | Sensitivity | Specificity (achieved) | ΔAUC vs PLOD3+FN1+TNC+LUM (P) |
| --- | --- | --- | --- | --- | --- | --- |
| PLOD3+FN1+TNC+LUM | IPMN vs PDAC | 56 vs 145 | 0.913 (0.839–0.989) | 0.481 | 0.921 | — |
| METTL7A+ISG15 | IPMN vs PDAC | 56 vs 145 | 0.945 (0.894–0.995) | 0.714 | 0.925 | +0.032 ($P = 0.322$) |
| FAT1+SEC11C | IPMN vs PDAC | 56 vs 145 | 0.911 (0.848–0.972) | 0.66 | 0.933 | -0.002 ($P = 0.940$) |
| CATB+EPIPL | IPMN vs PDAC | 56 vs 145 | 0.977 (0.952–0.999) | 0.92 | 0.915 | +0.064 ($P = 0.004$) |
| PLOD3+FN1+TNC+LUM | ND vs PDAC | 45 vs 145 | 0.992 (0.973–1.000) | 0.993 | 0.926 | — |
| METTL7A+ISG15 | ND vs PDAC | 45 vs 145 | 0.989 (0.971–1.000) | 0.979 | 0.92 | -0.003 ($P = 0.786$) |
| FAT1+SEC11C | ND vs PDAC | 45 vs 145 | 0.982 (0.944–1.000) | 0.933 | 0.913 | -0.010 ($P = 0.264$) |
| CATB+EPIPL | ND vs PDAC | 45 vs 145 | 1.000 (1.000–1.000) | 1 | 0.913 | +0.008 ($P = 0.264$) |

The mixed-direction pair METTL7A+ISG15 was constructed to test whether combining a protein that increases with one that decreases across the reference groups would yield more balanced detection. This was not borne out: the same-direction pair CATB+EPIPL achieved both higher sensitivity and comparable specificity. METTL7A+ISG15 therefore remains of interest for its biological rationale, pairing the loss of a normal ductal and metabolic protein with the gain of an interferon-stimulated stromal–immune protein, rather than for superior classification performance. FAT1+SEC11C, the only combination with confirmed, non-overlapping single-cell compartmental origins, was likewise supported by mechanistic interpretability rather than discriminative performance.

### 8. Identification of candidate diagnostic and therapeutic target proteins

Each of the four proteins selected through the unbiased screening pipeline discriminated PDAC from both normal comparators, with AUCs of 0.98–0.99 versus non-diseased pancreas and 0.93–0.97 versus normal-adjacent tissue (Figure 7A, B). Discrimination from precursor lesions was weaker and more variable for the individual proteins: against low- and high-grade IPMN, POSTN reached an AUC of 0.97 and ANXA2 0.93, whereas ISG15 and TIMP1 reached 0.87 and 0.84, respectively (Figure 8). Combined, the four proteins distinguished PDAC from IPMN with an AUC of 0.98 (95% CI 0.95–1.00; sensitivity 91% at 93% specificity), indicating that the panel captures complementary information not provided by any single member. This performance is notable because no protein was selected on the basis of discriminative performance; the panel was fixed by single-cell reproducibility, proteomic detection completeness, and extracellular accessibility before any ROC analysis. That all four proteins separated PDAC from normal tissue suggests that the selection criteria, particularly concordant marker status across independently collected single-cell cohorts, favored proteins with large and consistent tissue-level differences, although independent validation is required to exclude cohort-specific effects.

**Figure 7.**
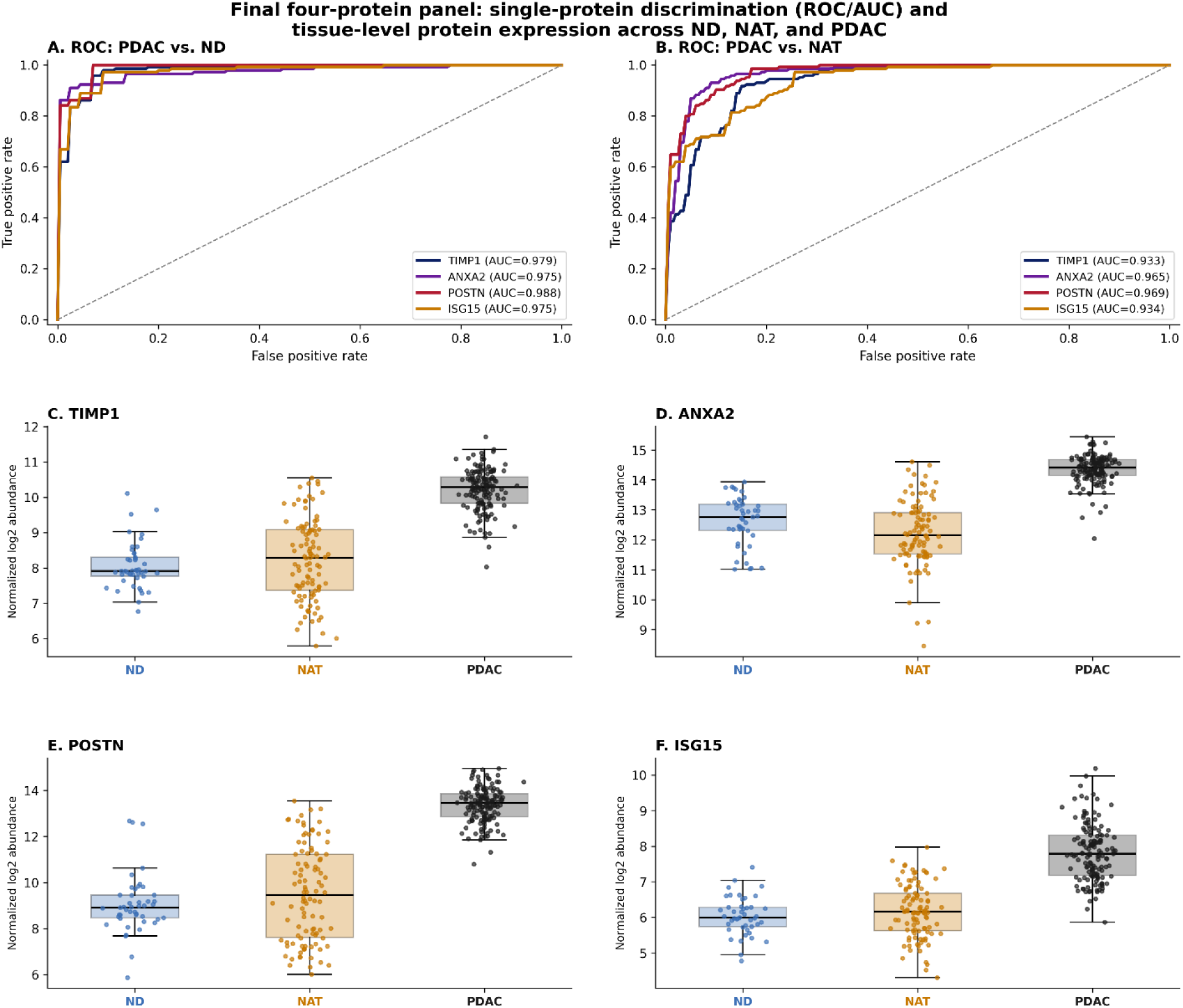
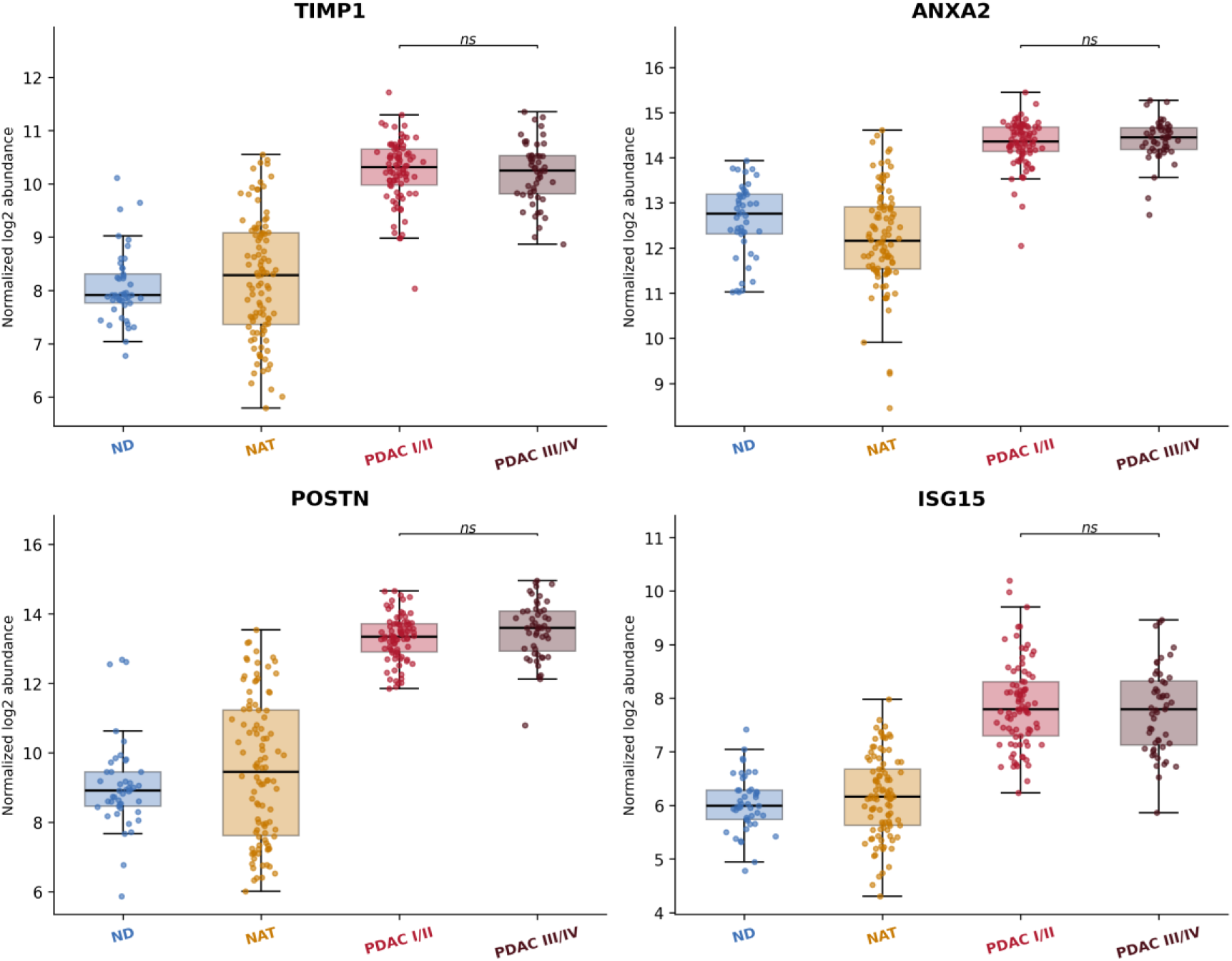
Final four-protein panel validation. (A, B) Single-protein ROC curves for TIMP1, ANXA2, POSTN, and ISG15 discriminating PDAC from ND (A) and PDAC from NAT (B), with bootstrap mean-curve AUC (n=500 resamples) shown for each protein. (C-F) Tissue-level protein abundance for each of the four proteins across ND, NAT, and PDAC; each point represents one patient sample, boxes show median and interquartile range. All four proteins were significantly elevated in PDAC relative to both ND and NAT (Mann-Whitney U test, Benjamini-Hochberg-adjusted P < 0.05 for both comparisons). Stage-resolved expression of the four-protein panel. (G-J) TIMP1, ANXA2, POSTN, and ISG15 abundance across ND, NAT, PDAC stage I/II, and PDAC stage III/IV. Each point represents one patient sample; boxes show median and interquartile range. None of the four proteins differed significantly between PDAC I/II and PDAC III/IV (ns, Mann-Whitney U test with Benjamini-Hochberg correction), indicating elevation is established by stage I/II.

**Figure 8.**
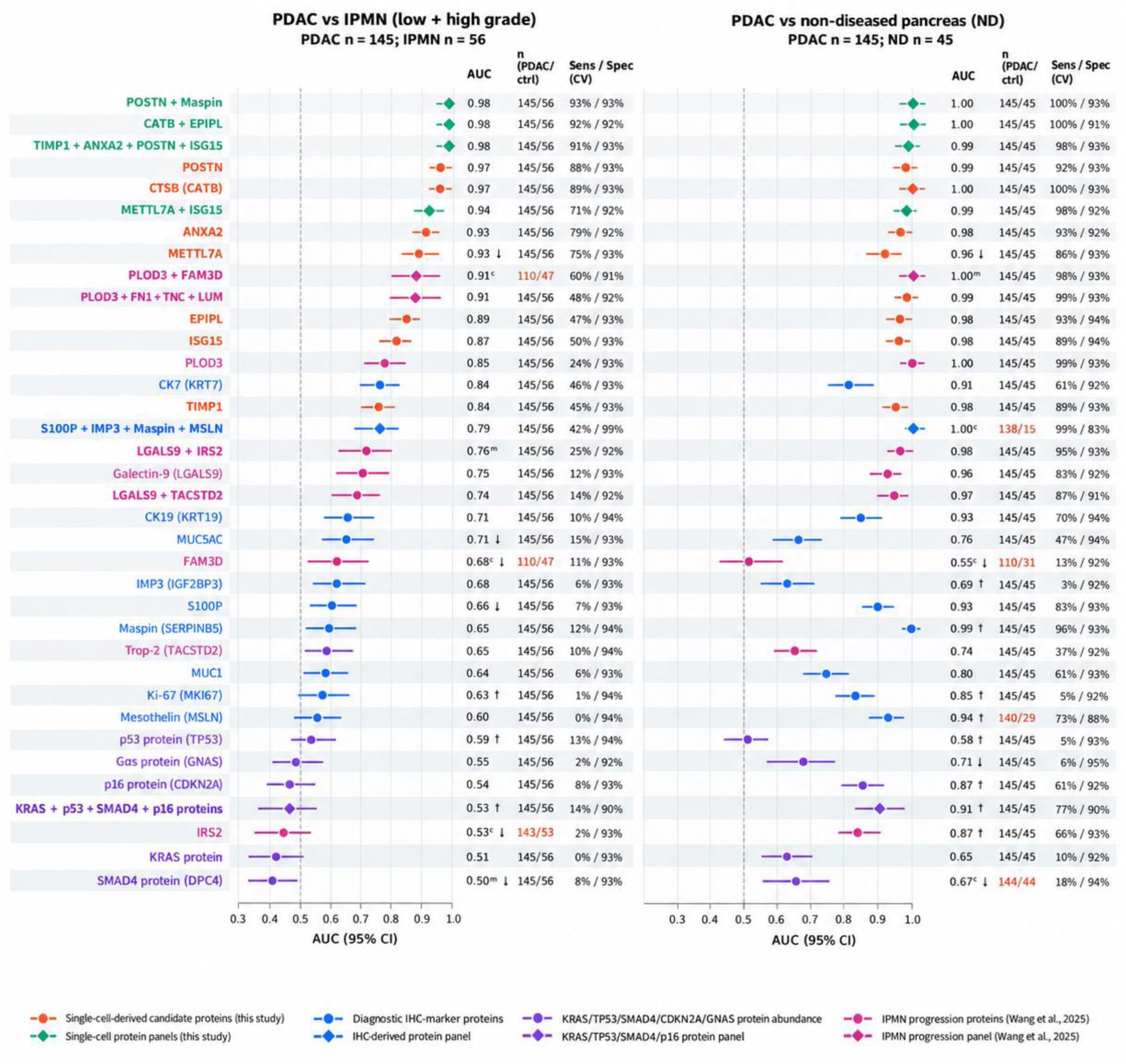
Tissue proteomic benchmarking of candidate panels against proteins associated with diagnostic IHC, molecular testing, and IPMN progression. AUC (95% CI) for PDAC (n = 145) versus low- and high-grade IPMN (n = 56, left) and non-diseased pancreas (n = 45, right). Circles, single proteins; diamonds, panels. All markers are tissue protein abundances measured by DIA-MS; KRAS, p53, SMAD4, and p16 do not represent mutation testing. Each row shows its best-performing missing-value approach: ᵐ median imputation; ᶜ complete-case (n in red); unmarked, below-detection imputation. †, AUC varied by >0.05 across approaches; ↓, lower in PDAC. Panels: cross-validated logistic regression. Sens/Spec at a training-fold 95%-specificity threshold.

TIMP1, ANXA2, and ISG15, by contrast, were each confirmed as significant markers of a tumor-enriched cell type in all four independent atlases and were detected in 99.7-100% of proteomic samples across every disease group with no concentration of missing measurements in any single comparator group. Among these three, ANXA2 showed the smallest difference in AUC between the ND and NAT comparisons (0.975 vs. 0.965, a gap of 0.010), compared with TIMP1 (0.979 vs. 0.933, a gap of 0.046) and ISG15 (0.975 vs. 0.934, a gap of 0.041). Because NAT tissue is anatomically adjacent to the tumor and may carry subclinical molecular changes despite appearing histologically normal, it represents a more conservative and clinically realistic comparator than distant non-diseased pancreas. ANXA2’s comparatively stable performance across both comparators suggests its discriminative signal is less sensitive to which normal tissue type is used as the reference, a property that may be advantageous if the panel is ultimately validated against normal-adjacent rather than fully non-diseased tissue in a clinical setting.

The tissue-level expression data (Figure 7C–F) further differentiate the four proteins by the consistency, rather than only the magnitude, of their elevation in PDAC. TIMP1 and ANXA2 showed the tightest within-group spread in the PDAC cohort (coefficient of variation 5.8% and 3.3%, respectively), indicating that their elevation is broadly uniform across patients rather than concentrated in a subset. ISG15 and POSTN showed comparatively wider PDAC-group spread, and POSTN’s NAT-group distribution was visibly bimodal, with one subset of NAT samples resembling ND-level abundance and another approaching PDAC-level abundance. This bimodality in NAT is consistent with the possibility that some normal-adjacent samples in this cohort already carry field-effect molecular changes for POSTN specifically, and warrants examination of whether these samples share other clinical or pathological features before POSTN is advanced as a standalone diagnostic marker.

Because early detection is the primary clinical objective, we further examined whether these proteins were elevated specifically in early-stage disease rather than only in advanced PDAC. Disaggregating the PDAC group into stage I/II (n=85) and stage III/IV (n=51) (the remaining 9 of the 145 PDAC cases were not assigned to either subgroup) revealed no significant difference between these two subgroups for any of the four proteins (TIMP1, ANXA2, POSTN, ISG15; Mann-Whitney U test, Benjamini-Hochberg-adjusted P>0.05 for all comparisons; Figure 7G-J), indicating that the elevation of these candidates over non-diseased and normal-adjacent tissue is already fully established at stage I/II and does not further increase with disease progression to stage III/IV. Taken together, these tissue-level results support two complementary priorities for future validation: TIMP1 and ANXA2 as the more uniformly and reproducibly elevated markers, warranting priority evaluation as candidates for a blood-based single-cutoff diagnostic test. ANXA2 is especially attractive because surface-localised ANXA2 drives invasion and metastasis in PDAC and is blocked by antibody ^61^, it serves as a vaccine-identified antigenic target ^62^, its intracellular pool mediates gemcitabine resistance through NF-κB ^63^, and high expression independently predicts worse survival ^64^. TIMP1 is a well-established, cross-validated biomarker of pancreatic and other epithelial malignancies: elevated serum and tissue levels reliably distinguish cancer from both healthy and benign disease states and improve diagnostic and prognostic accuracy when combined into multi-marker panels, though its prognostic value can be confounded by jaundice and cachexia^65–68^. Mechanistically, TIMP1 transcription is directly induced by oncogenic K-Ras through ERK2 signaling and is secreted predominantly by stromal cells, including cancer-associated fibroblasts and mesenchymal stem cells, to drive tumor cell proliferation, venous invasion, and disease progression via PI3K/AKT, ERK1/2, and cMet–RAP1 signaling across multiple cancer types^65,69,70^. In our single-cell analysis, TIMP1 was enriched predominantly in fibroblasts, with additional expression in macrophages, linking its tissue-level increase to the stromal and myeloid remodeling observed in PDAC. POSTN as the single strongest discriminator by magnitude, better suited to a role emphasizing maximal separation where subpopulation-level heterogeneity is an acceptable trade-off This heterogeneity itself is mechanistically grounded: POSTN is secreted almost exclusively by activated stellate cells and cancer-associated fibroblasts in an autocrine loop that sustains stromal activation, confers chemoresistance, and shortens survival (Erkan et al., 2007), and single-cell and spatial transcriptomic studies show that only a specific ECM-remodeling fibroblast subpopulation drives this program through direct POSTN–ITGAV/ITGB5 engagement of tumor cells, activating PI3K/AKT/β-catenin signaling and epithelial–mesenchymal transition while also promoting angiogenesis via Erk/VEGF signaling^71–73^. Consistent with a non-uniform, immunologically consequential source, POSTN⁺ fibroblasts spatially and transcriptionally co-localize with immunosuppressive SPP1⁺ macrophages across multiple independent cohorts and correlate with worse overall survival^72^, and elevated POSTN is independently associated with increased M2 macrophage infiltration and reduced predicted responsiveness to checkpoint immunotherapy^74,75^. ISG15 occupies an intermediate position, combining full single-cell reproducibility with a secreted extracellular form^76^, and direct functional involvement in PDAC, where macrophage-derived free ISG15 sustains cancer stem cell self-renewal ^77^, ISGylation maintains their metabolic plasticity ^54^, and pathway knockdown lowers PD-L1 and improves anti-PD-1 efficacy, and pathway knockdown lowers PD-L1 and improves anti-PD-1 efficacy ^78^ that additionally support its use as a treatment-monitoring or delivery-relevant target rather than a purely diagnostic one.

### 9. Candidate panels outperform proteins associated with diagnostic IHC, molecular testing, and IPMN progression in distinguishing PDAC from IPMN

To place the candidates in a clinical context, we benchmarked them against proteins corresponding to diagnostic IHC markers, the protein products of genes assessed in sequencing-based molecular testing, and the IPMN-progression proteins and combinations reported by Wang et al.²⁹ (Figure 8). Against low- and high-grade IPMN (145 versus 56 samples), three candidate panels reached an AUC of 0.98: POSTN+maspin (sensitivity 93% at 93% specificity), CATB+EPIPL (92% at 92%), and TIMP1+ANXA2+POSTN+ISG15 (91% at 93%), and POSTN and CTSB alone reached 0.97. Proteins corresponding to IHC markers discriminated less well. CK7 was the most discriminative (AUC 0.84), the others ranged from 0.60 to 0.71, and S100P and MUC5AC were more abundant in IPMN than in PDAC; combined, S100P+IMP3+maspin+MSLN reached 0.79 (42% at 89%). The refitted IPMN-progression combinations reached 0.74–0.91, the highest being PLOD3+FN1+TNC+LUM (48% at 92%) and PLOD3+FAM3D in complete-case analysis (110 versus 47 samples). In paired comparisons, CATB+EPIPL, TIMP1+ANXA2+POSTN+ISG15, POSTN+maspin, POSTN, and CTSB each exceeded the IHC-associated panel (ΔAUC 0.18–0.19, P < 0.001) and PLOD3+FN1+TNC+LUM (ΔAUC 0.05–0.07, P ≤ 0.014). The protein products of KRAS, TP53, SMAD4, and CDKN2A did not discriminate PDAC from IPMN, either individually (AUC 0.50–0.59) or combined (0.53, 95% CI 0.37–0.63); this result concerns protein abundance only and does not evaluate sequencing-based mutation tests.

Against non-diseased pancreas (145 versus 45 samples), candidate and comparator panels performed similarly. Candidate panels reached AUCs of 0.99–1.00, refitted IPMN-progression combinations 0.97–1.00, and the IHC-associated panel 1.00 in complete-case analysis (138 versus 15 samples); differences between the leading panels were small (|ΔAUC| ≤ 0.01). Several IHC-associated proteins also discriminated well individually, including maspin (0.99), MSLN (0.94), S100P (0.93), and CK19 (0.93). However, maspin, MSLN, Ki-67, IMP3, p16, and IRS2 were detected less often in non-diseased pancreas than in PDAC, and their AUCs changed by more than 0.05 depending on how missing values were handled (Supplementary Figure S11). Maspin, for example, fell from 0.99 to 0.72 under median imputation. The candidate proteins were detected in at least 98% of samples in every group and were unaffected. The advantage of the candidate panels therefore lies specifically in distinguishing invasive carcinoma from precursor lesions, and it persisted when each comparator was evaluated under its most favorable missing-value approach.

## DISCUSSION

A methodological choice in this study warrants explicit justification: why we used the pancreatic neoplasia disease-spectrum as an ordered proteomic reference framework, rather than a literal patient-level progression sequence, and why we selected proteins by a stepwise, monotonic-ordering criterion rather than a conventional two-group comparison.

Because IPMN progresses through a well-defined, clinically established sequence of dysplasia grades culminating in invasive carcinoma, we used this progression primarily as an ordered proteomic reference framework for testing directional consistency in candidate PDAC biomarkers, rather than as a literal patient-level temporal sequence or a study subject in its own right: our single-cell analyses were restricted to PDAC tissue, and no single-cell sequencing of IPMN specimens was performed in this work. This structure, comprising non-diseased pancreas, normal-adjacent tissue, low-grade IPMN, high-grade IPMN, invasive IPMN, and PDAC, provided an ordered proteomic scaffold against which a candidate marker’s consistency of directional change, and therefore its suitability for specific and early PDAC detection, could be tested directly, in contrast to single-timepoint case-control marker discovery. We emphasize that normal-adjacent tissue is not a progression stage and that PDAC is not the obligatory clinical outcome of invasive IPMN; the ordering reflects an analytical reference structure for biomarker screening across the pancreatic neoplasia spectrum, not a claim about individual patients’ disease course. This reference framework is complicated, however, by the known biological heterogeneity of IPMN precursor lesions even within a single clinical grade^79,80^, heterogeneity we independently confirmed at the proteomic level through bootstrap-supported dispersion analysis. Reflecting this heterogeneity, different groups have proposed distinct biomarker panels tailored to specific pairwise comparisons rather than the full spectrum^81–83^. To distinguish these histopathologic reference groups robustly despite this heterogeneity, rather than fitting a panel to a single pairwise comparison, we applied a stringent monotonic, stepwise ordering criterion requiring consistent directional change across all six reference groups, and additionally incorporated downregulated proteins alongside the upregulated markers emphasized in most prior panels. We reasoned that this combined approach would increase specificity for early PDAC detection and reduce sensitivity to fluctuating protein signals that a non-monotonic, single-direction marker set might otherwise introduce.

The single-cell component of this study sits within a rapidly maturing literature in which single-cell profiling of PDAC has moved from cataloguing cell types toward defining spatially and clinically resolved cell states. Chen and colleagues combined single-cell, single-nucleus, and spatial transcriptomics in PDAC tissues stratified by invasion of nerves and vessels status and showed that invaded nerves are surrounded by NLRP3+ tumor-associated macrophages and myofibroblastic CAFs, whereas non-invaded nerves associate with tertiary lymphoid structures, and that a TGFBI+ Schwann cell subset actively promotes invasion of nerves and vessels^15^. Kanu and colleagues extended profiling beyond the primary tumor and showed that macrophages from PDAC tumors are polarized toward an immunosuppressive M2 phenotype, with upregulation of APOE, CD163, WNT5A, THBS1, SPP1, and HIF1A, whereas macrophages from the premetastatic liver of the same patients retain a distinct, M1-skewed profile^33^. Two features of these studies bear directly on the present analysis. First, the macrophage genes they highlight overlap closely with the immunoregulatory and hypoxia-adaptation modules identified here, supporting the reproducibility of the C1QA+ TAM program across independent cohorts. Second, both studies underscore that tissue of origin determines cell state: because three of the four atlases integrated here contributed peripheral blood in addition to tumor tissue, blood-derived and tissue-derived cells of the same lineage should be distinguished before compositional or marker-based inferences are drawn (Supplementary Figure S7).

The CAF-associated candidates identified here can be placed within the consensus framework that has emerged from single-cell studies of fibroblasts across organs. Lavie and colleagues proposed that CAFs be classified hierarchically into myofibroblastic/ECM-remodeling, inflammatory/immune-regulatory, and antigen-presenting programs, with ACTA2, TAGLN, and POSTN marking myofibroblastic CAFs, IL6 and CXCL12 marking inflammatory CAFs, and CD74 and HLA class II genes marking antigen-presenting CAFs, and noted that THBS2 is among the markers of ECM-remodeling CAFs in both murine and human tumors^58^. The stepwise-increasing proteins attributed to fibroblasts in our analysis-SFRP2, THBS2, IGFBP5, TIMP1, and CRTAP-belong to the matrix-remodeling rather than to the inflammatory or antigen-presenting arm of this classification, consistent with their monotonic increase across a progressively desmoplastic reference sequence and supporting their interpretation as markers of cumulative stromal activation rather than of fluctuating inflammatory states. This distinction is therapeutically relevant, because CAF-directed strategies increasingly aim to reprogram rather than to deplete the stroma: indiscriminate depletion of myofibroblastic CAFs can accelerate disease, whereas normalizing CAFs, or exploiting the peritumoral CAF barrier itself as a drug depot, has been proposed as a means of improving delivery of chemotherapy and immunotherapy in PDAC^58,59^.

For the macrophage compartment, our module-level analysis indicates that the genes restricted to macrophages are predominantly effectors of the immunoregulatory program (C1QA, C1QB, MS4A4A, MSR1, OLR1, CD163, and FCGR2A), whereas several ligands and mediators commonly described as TAM-associated (CSF1, CXCR4, TGFB1, SPP1, and VEGFA) are preferentially expressed by other stromal and immune populations. This division has implications for TAM-directed therapy. Strategies aimed at macrophage-associated receptors and signaling axes, including CSF1-CSF1R and CCL2-CCR2 blockade, have shown limited single-agent clinical activity, and attention has shifted toward reprogramming macrophages toward an immunostimulatory state through epigenetic and metabolic regulators, cytokine- and cell-based approaches, and macrophage-targeted nanoplatforms^60^. If the ligands that sustain the TAM phenotype are supplied largely by non-macrophage compartments, as our data suggest, receptor blockade confined to macrophages may be readily bypassed, and markers shared between the fibroblast and macrophage compartments-TIMP1, ISG15, and CTSB in our analysis-may reflect the combined stromal-myeloid program more faithfully than markers of either cell type alone. Finally, both compartments are plastic under treatment: chemotherapy has been shown to reprogram PDAC CAFs toward a neural, axonal-guidance phenotype (NRP1, NEGR1, SLIT2, and EPHA3) that is enriched near sites of periinvasion of nerves and vessels and further enriched in metastatic recurrence^19^. Because all specimens analyzed here were treatment-naive, our findings describe the untreated microenvironment, and the stability of the nominated markers under chemotherapy remains to be tested.

The macrophage results merit two further comments. First, the genes that were restricted to macrophages in our atlas are effectors of an established phagocytic and immunoregulatory program-complement (C1QA, C1QB), scavenger receptors (MSR1, OLR1, CD163), and Fc receptors (FCGR2A)-whereas the corresponding ligands and upstream mediators were assigned to other compartments: CSF1 to mast cells, CXCR4 to lymphoid and dendritic cells, TGFB1 to platelets and lymphocytes, and VEGFA to mast cells. Read together with the paracrine architecture described in the CAF analysis above, this suggests that the C1QA+ TAM state in PDAC is maintained by inputs distributed across the microenvironment rather than generated cell-autonomously, which is consistent with the limited single-agent activity of therapies directed at individual macrophage receptors and with the current emphasis on reprogramming macrophages instead of depleting them^60^. Two assignments also have straightforward biological explanations and should not be over-read: FCGR3A was more enriched in NK cells than in macrophages because it encodes CD16, and CD74 with HLA class II genes was shared across macrophages, dendritic cells, and B cells because all three are professional antigen-presenting populations, a program that PDAC fibroblasts can also acquire^12,58^. Second, SPP1 was not identified as a macrophage marker in our analysis, although SPP1+ macrophages are among the best-described tumor-associated macrophage states in PDAC and across human cancers, where the CXCL9:SPP1 axis defines macrophage polarity and tracks with outcome, and where tumor macrophages upregulate SPP1 relative to macrophages from unaffected tissue^25,33^. The discrepancy most likely reflects the test rather than the biology: marker detection asks whether a gene is enriched in one cell type relative to all others, so broadly expressed genes such as SPP1 can be missed even when they are abundantly expressed in macrophages. Confirming the SPP1+ state in this atlas would require a within-compartment comparison, for example a pseudobulk contrast of tumor versus adjacent-normal macrophages, which was not performed here.

The depletion of dendritic cells was the largest compositional change we observed, and it is supported by both experimental and clinical evidence in PDAC. In mouse models, a paucity of conventional dendritic cells produces dysfunctional immune surveillance and favours tumour-promoting Th17 responses, while restoring conventional dendritic cells restrains early disease and improves responsiveness to radiation therapy^49^. In patients, the deficit is not confined to the tumour: circulating myeloid and plasmacytoid dendritic cells are reduced and show increased apoptosis in PDAC, these changes are only partially reversed after tumour resection, and patients surviving more than two years retain significantly higher blood dendritic cell levels than those surviving less than one year ^84^. Notably, comparable reductions were seen in chronic pancreatitis in that study, indicating that the dendritic cell deficit tracks with pancreatic inflammation rather than being specific to malignancy^84^. Our observation of a selective loss of a CLEC4C+/LILRA4+ plasmacytoid compartment is consistent with this picture and extends it to the tissue level, but two cautions apply. First, because plasmacytoid dendritic cells are rare and particularly vulnerable to dissociation-related dropout, the magnitude of the depletion we report should be regarded as an upper bound until confirmed by an orthogonal method such as immunohistochemistry or flow cytometry. Second, since circulating dendritic cell numbers are themselves reduced in PDAC, any comparison that mixes blood-derived with tissue-derived cells will confound the systemic and local components of this deficit, which is a further reason to keep the two sources separate (Supplementary Figure S7). If confirmed, the combination of a systemic and an intratumoural dendritic cell deficit would strengthen the rationale for dendritic cell-restoring strategies, such as FLT3L and CD40 agonism, in this disease^49^.

Placed against the wider PDAC literature, the fibroblast programs detected here-ECM-receptor interaction, focal adhesion, integrin signaling, and PI3K-Akt-are the transcriptional counterpart of the desmoplastic reaction that defines the histology of this tumour, which single-cell studies have resolved into at least three recurrent states: myofibroblastic CAFs adjacent to tumour cells, inflammatory CAFs secreting IL-6 and related mediators, and antigen-presenting CAFs expressing MHC class II^12,55,58,85^. Interpreting stromal enrichment as uniformly tumour-promoting would nonetheless be a mistake. Genetic depletion of alpha-SMA-positive myofibroblasts and deletion of Sonic hedgehog both accelerate disease and shorten survival in mouse PDAC^86,87^, and pharmacological hedgehog inhibition shifts CAF composition toward the inflammatory state rather than removing the stroma^88^. What separates tractable from counterproductive stromal intervention appears to be the targeting of signaling rather than of cells: focal adhesion kinase inhibition reduces fibrosis and sensitizes tumours to checkpoint blockade^45^, beta-1 integrin and STAT3 mechanosignaling can be interrupted without ablating the matrix^46^, and TGF-beta-driven LRRC15+ myofibroblasts identify patients unlikely to respond to anti-PD-L1 therapy^56^. Our compartment-resolved analysis adds a practical qualification to this literature: the same integrin and PI3K-Akt programs are expressed by endothelial and myeloid cells as well as fibroblasts, so they are unlikely to be silenced by fibroblast-directed therapy alone. A recent mechanistic study illustrates how such a program can be instructed from outside the fibroblast compartment: in PDAC, neutrophil extracellular traps engage integrin alpha-5-beta-1 (ITGA5) on fibroblasts through NET-DNA, initiating a FAK/SRC-YAP-IL-6-JAK/STAT3 autocrine cascade that specifies inflammatory CAFs and promotes CD8+ T-cell exhaustion, with tumour-derived NGAL induced by IL-17 sustaining NETosis upstream; blocking ITGA5 or the IL-17 axis shifts fibroblasts back toward a myofibroblastic state and synergises with PD-1 blockade^22^. This places the integrin and STAT3 components detected in our fibroblast compartment downstream of a defined and druggable extracellular input, and it also exposes a gap in the present atlas: neutrophils were not recovered as an annotated population in the integrated datasets, so a NET-driven axis could not be evaluated here and would require cohorts in which granulocytes are preserved. Following the same logic, CAF-directed nanomedicine has moved away from non-specific fibroblast depletion toward precision stromal modulation, using carriers engineered to match the physical and spatial properties of CAF-rich stroma^21,59^.

The myeloid findings fit an equally consistent body of work. Complement-high monocytes and macrophages have been described in both the blood and tumours of patients with PDAC^89^, interleukin-1-beta-producing macrophages sustain a pathogenic inflammatory circuit^30^, SPP1 and CXCL9 define opposing macrophage polarities that track with outcome across human cancers^25^, and a fraction of pancreatic tumour macrophages derives from embryonic haematopoiesis and expands locally to drive fibrosis^31^. The density and spatial organization of myeloid infiltration carry prognostic weight in resected PDAC^32,90^, which is consistent with our observation that macrophages were the most transcriptionally distinct immune population in the integrated atlas. Considered alongside the checkpoint-engaged T-cell profile and the dendritic cell deficit described above, these features describe the immune-excluded phenotype that characterizes most pancreatic tumours: the spatial distribution of intratumoural T cells, rather than their overall number, correlates with survival^91^; stromal CXCL12 forms a coating on carcinoma cells that impedes T-cell attack^44^; CXCR4 inhibition elicits an integrated immune response in patients with pancreatic and colorectal cancer^92^; and the quality rather than the quantity of neoantigens distinguishes long-term survivors^93^. That several mediators of this phenotype, including CSF1, CXCR4, and TGFB1, were assigned to non-macrophage compartments in our data reinforces the view that immune exclusion in PDAC is a property of the tissue rather than of any single cell type.

Two methodological points follow from this synthesis. First, our analysis is cross-sectional and treatment-naive, and therefore describes a baseline state. Spatially resolved and multimodal studies have shown that the same compartments are reorganized into discrete niches^26,94^ and are substantially remodelled by chemotherapy, with loss of myofibroblastic CAFs, gain of inflammatory and neural CAF programs, and altered myeloid composition after treatment^19,20,23^. The programs reported here should therefore be regarded as the untreated reference against which such changes are measured, not as stable features of the disease. Second, the single-cell signature scores should be read conservatively. The KRAS, MAPK, mTOR, and PI3K gene sets used here (Supplementary Table S4) contain many inflammatory, metabolic, and cytoskeletal genes that are broadly expressed by myeloid and stromal cells, so a high score in macrophages indicates expression of pathway member genes rather than evidence of RAS pathway output or of oncogenic mutation in those cells; a per-compartment pseudobulk contrast across the disease spectrum would test that inference directly and was not performed here. With those caveats, the value of defining these programs is that they constrain how the proteomic candidates are interpreted: the stepwise proteins nominated in this study map onto the fibroblast program (THBS2, SFRP2, IGFBP5, TIMP1) and the myeloid program (CTSB, TYMP, CTSS), so their behaviour across the disease spectrum is most plausibly read as a readout of coordinated stromal-immune remodeling rather than of tumour-cell content alone.

### Limitations

Several limitations of the present analysis should be noted, some of which cannot be resolved with the data currently available and would require additional experiments or cohorts. First, tumor-enriched and tumor-depleted cell populations were identified using Fisher’s exact test on pooled cell counts across all patients within each condition; this approach does not account for the patient as the unit of biological replication and is susceptible to pseudoreplication, whereby differences driven by a small number of patients with high cell yield could be mistaken for a population-level effect. Per-sample cell-type proportions and a mixed-effects or compositional statistical framework, adjusting for patient, batch, and cellularity, would be a more rigorous approach; although per-sample proportions are reported descriptively (Figure 2A and 2C), such a model was not fitted in the present study. Second, the coordinated CAF-TAM-pDC remodeling program we describe is inferred from co-enrichment and depletion patterns and from pathway hubs that are common to many inflamed and desmoplastic tumor microenvironments; we did not perform ligand-receptor inference (e.g., CellPhoneDB, NicheNet, or LIANA) or spatial co-localization analysis to establish direct sender-receiver signaling relationships or physical proximity among these populations, and the term should be read as describing a coordinated pattern rather than a mechanistically or causally established signaling circuit. Third, pDC depletion is based on a small number of canonical markers (CLEC4C, LILRA4) in a rare population that is particularly sensitive to dissociation-related dropout; we did not perform additional doublet-filtering or marker-panel sensitivity analyses to confirm per-patient reproducibility of this finding. Fourth, the KEGG pathway enrichment analysis ranked genes by cluster-marker status relative to all other cell types, which captures lineage identity rather than within-lineage activation differences across disease stages; a pseudobulk, per-patient contrast across the ND-to-PDAC spectrum within each cell type would more directly test the progression hypothesis and was not performed here. Fifth, proteomic panel discrimination was evaluated using bootstrap resampling on the same discovery cohort used for feature selection; while this provides a stable estimate of the panel’s performance and its uncertainty on the available data, it does not constitute independent or external validation, and an held-out or externally collected cohort was not available for this analysis. Sixth, the published four-protein comparator panel was re-fit on our own cohort using the same logistic-regression procedure applied to our two-protein panels, rather than applying its originally published coefficients; we consider this the more consistent comparison given differences in cohort and platform but note it as a methodological choice that should be made explicit. Seventh, we have not evaluated the specificity of TIMP1, ANXA2, POSTN, or ISG15 against non-PDAC conditions such as pancreatitis or other gastrointestinal malignancies, nor have we measured any of these proteins in serum or plasma; the tissue-level associations reported here should be regarded as candidates for, rather than evidence of, blood-based detection or targeted delivery, both of which require dedicated functional and clinical studies beyond the scope of this work. Finally, potential differences in collection context, systemic inflammation, or tissue composition between non-diseased pancreas and normal-adjacent tissue were not directly modeled and could contribute to some of the differences we attribute to disease status.

Several additional limitations warrant explicit statement. The single-cell cohort comprises only 26 patients unevenly distributed across four atlases (14, 6, 3, and 3), and leave-one-dataset-out or per-patient subsampling analyses were not performed to exclude dominance of the largest cohort. In the proteomic data, specimens were restricted to >15% neoplastic cellularity, but residual variation in stromal and immune proportions was not adjusted for, and the contribution of acquisition batch or processing site to the monotonic trends was not formally tested. Panels were evaluated by L2-regularized logistic regression in repeated cross-validation, with imputation and scaling fitted within training folds, refitted bootstrap confidence intervals, and a fold-derived 95%-specificity threshold; specificity in held-out samples was lower (approximately 91–94%), calibration was not assessed, and cross-validation within a single cohort does not substitute for external validation. The benchmarking has three further constraints. First, several reference proteins, including maspin, CDKN2A, Ki-67, and MSLN, were detected far less often in non-diseased pancreas than in PDAC, so their performance against non-diseased tissue depended on the missing-value assumption (Supplementary Figure S11). Second, each comparator was reported under its best-performing missing-value approach, which favors the reference markers but adds optimism to their estimates. Third, all markers were measured as tissue protein abundance: IHC-associated markers were not evaluated by staining, and KRAS, p53, SMAD4, and p16 protein abundance is not equivalent to sequencing-based mutation testing. CA19-9 could not be benchmarked because it is a carbohydrate epitope on mucin backbones rather than a discrete protein, and measuring it requires glycoproteomic or lectin-based assays not included in this global proteomic dataset. Finally, no leave-one-group-out analysis was performed for the stepwise selection criterion, so removing the small invasive-IPMN group (n = 8) or another reference group could alter which proteins satisfy monotonicity.

### Conclusion

By integrating single-cell transcriptomic atlases with an ordered tissue proteomic reference framework spanning the pancreatic neoplasia spectrum, we linked the stromal and myeloid remodeling of PDAC to protein-level markers in tissue. The resulting panels, CATB+EPIPL and the stromal–immune panel TIMP1+ANXA2+POSTN+ISG15, distinguished PDAC from low- and high-grade IPMN more accurately than protein-abundance models of established diagnostic IHC markers and refitted published IPMN-progression combinations. Against non-diseased pancreas, they performed comparably to those comparators. These findings are derived from a single cohort and from mass spectrometry–based protein abundance, and they nominate rather than establish clinical markers. Independent validation, immunohistochemical evaluation in EUS-guided fine-needle biopsy specimens, and measurement in blood will be required to determine their diagnostic utility.

## Data Availability

All data generated in this study are available from the corresponding author upon reasonable request.

## Acknowledgements

C.M.S., T.T., and J.F.F. were supported by the National Cancer Institute (NCI) of the National Institutes of Health under award number U01CA239522. This publication was supported, in part, by the Indiana Clinical and Translational Sciences Institute, funded in part by Grant Number UM1TR004402 from the National Institutes of Health, National Center for Advancing Translational Sciences, Clinical and Translational Sciences Award. Support for T.T. and C.M.S was provided under this award. The content is solely the responsibility of the authors and does not necessarily represent the official views of the National Institutes of Health. The authors thank Dr. Ralph Hruban and Yuefan Wang for their insightful feedback and for providing the proteomic data used in this study.

## Supplementary Materials

### Supplementary Tables

**Supplementary Table S1.** Clinical and data information for the integrated scRNA-seq cohort. Patient-level summary of the 26 patients by clinical stage and source dataset with the number of QC-passed cells per patient (324,662 cells in total); clinical and sample information for each dataset, including tissue source and pre- and post-QC gene and cell numbers and quality-control thresholds; and data sources and original publications.

**Supplementary Table S2.** Cell-type marker genes in the integrated atlas. Seurat FindAllMarkers results (Wilcoxon rank-sum test) for each of the 12 annotated cell types relative to all other cells, with average log2 fold change, fractions of expressing cells within (pct.1) and outside (pct.2) the cell type, and nominal and Bonferroni-adjusted P values.

**Supplementary Table S3.** KEGG pathway enrichment and oncogenic signature comparisons across cell types. Cell-type-specific gseKEGG results (set size, enrichment score, normalized enrichment score, adjusted P value, and core-enrichment genes) and pairwise Wilcoxon rank-sum comparisons of KRAS, MAPK, mTOR, and PI3K signature scores between cell types.

**Supplementary Table S4.** Gene signatures used for oncogenic pathway scoring. Member genes and MSigDB source gene sets for the KRAS, MAPK, mTOR, and PI3K signatures.

### Supplementary Figures

**Supplementary Figure S1.**
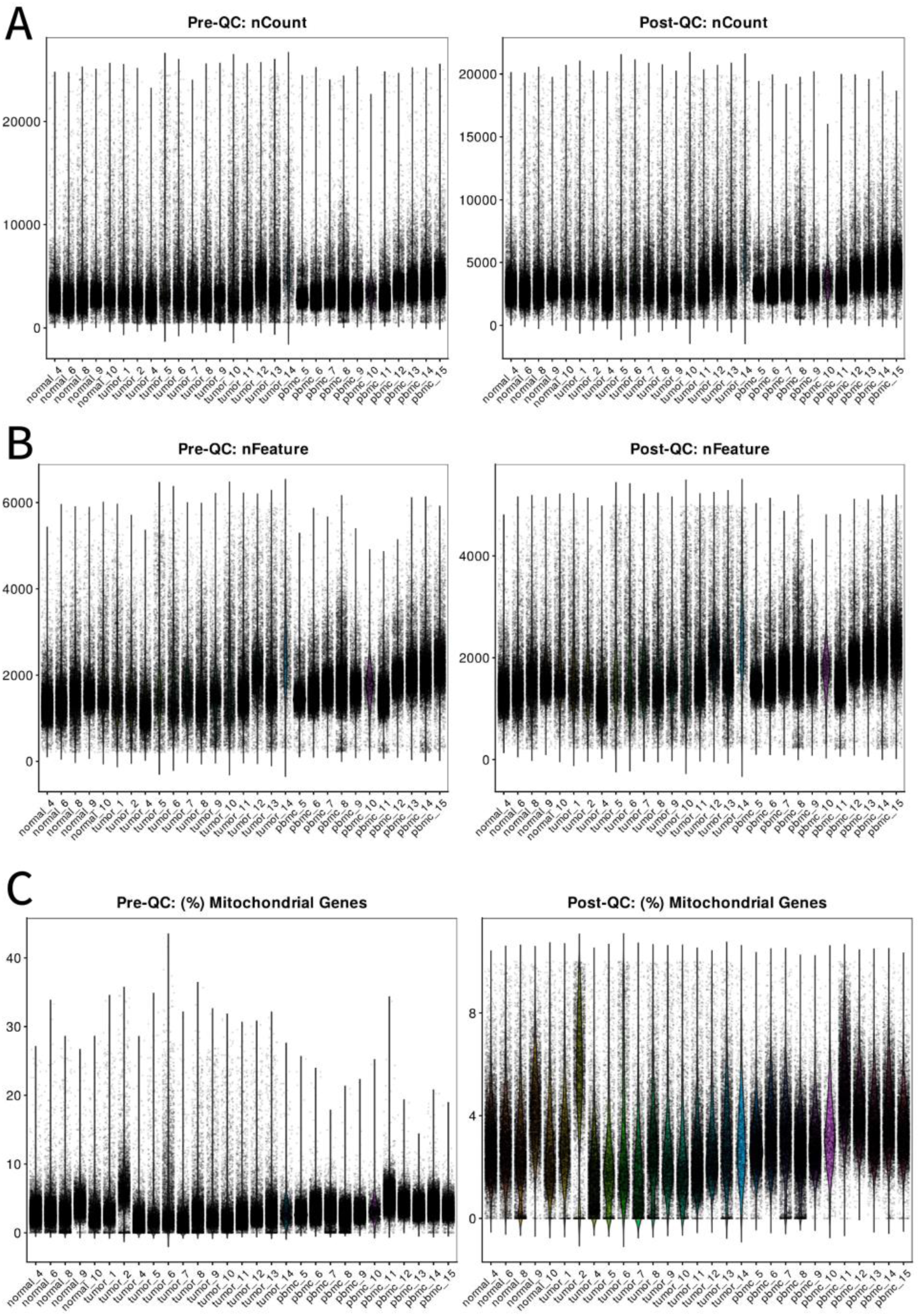
Quality control of GSE278694. Violin plots of (A) total UMI counts per cell (nCount), (B) number of detected genes per cell (nFeature), and (C) percentage of mitochondrial transcripts per cell for each sample before (left) and after (right) quality control. Samples include adjacent normal tissue, tumor, and PBMC. Cells with fewer than 200 or more than 5,000 detected genes, more than 20,000 UMIs, or more than 10% mitochondrial transcripts were removed.

**Supplementary Figure S2.**
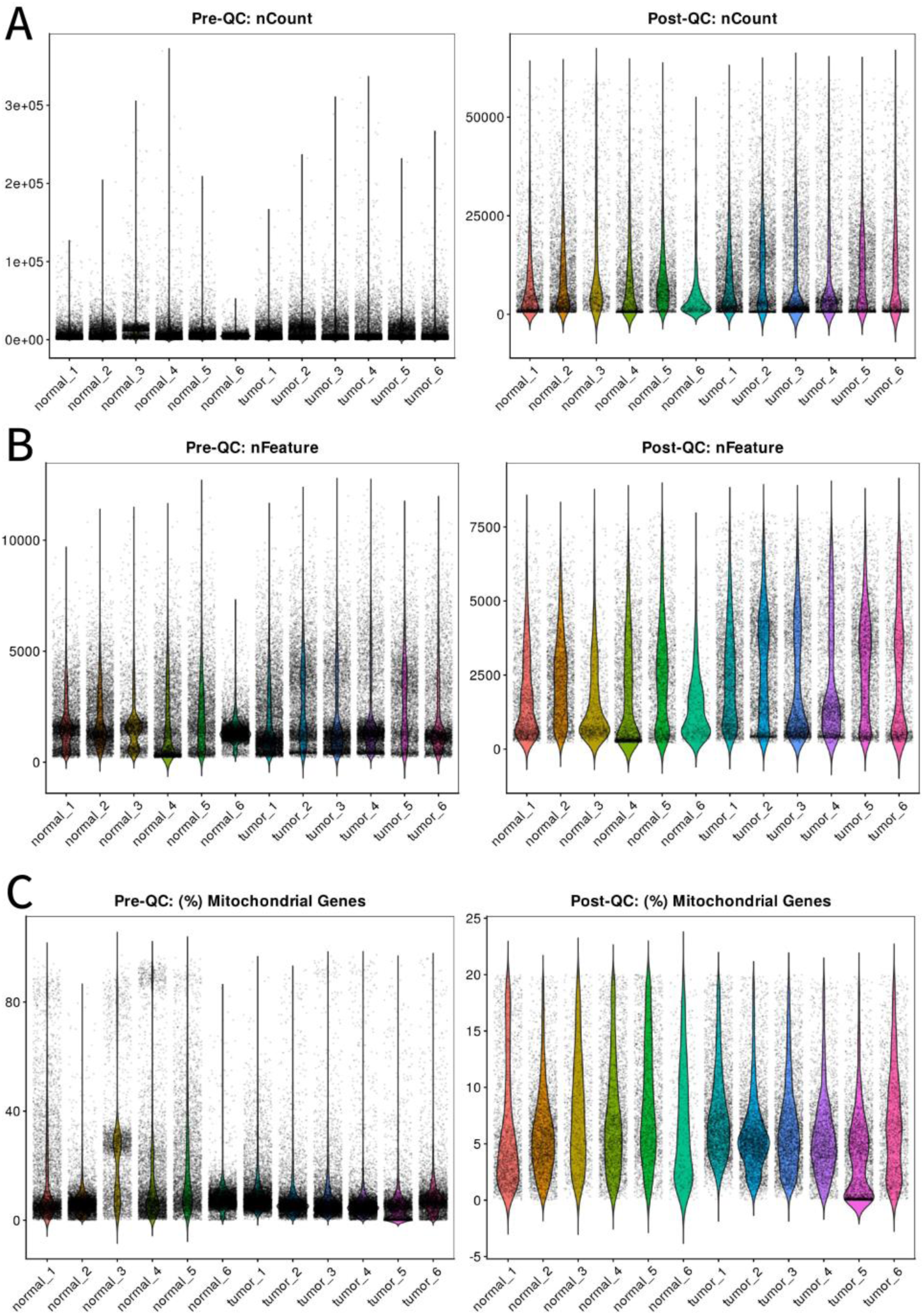
Quality control of GSE212966. Violin plots of (A) total UMI counts per cell (nCount), (B) number of detected genes per cell (nFeature), and (C) percentage of mitochondrial transcripts per cell for each sample before (left) and after (right) quality control. Samples include adjacent normal tissue and tumor. Cells with fewer than 200 or more than 8,000 detected genes, more than 60,000 UMIs, or more than 20% mitochondrial transcripts were removed.

**Supplementary Figure S3.**
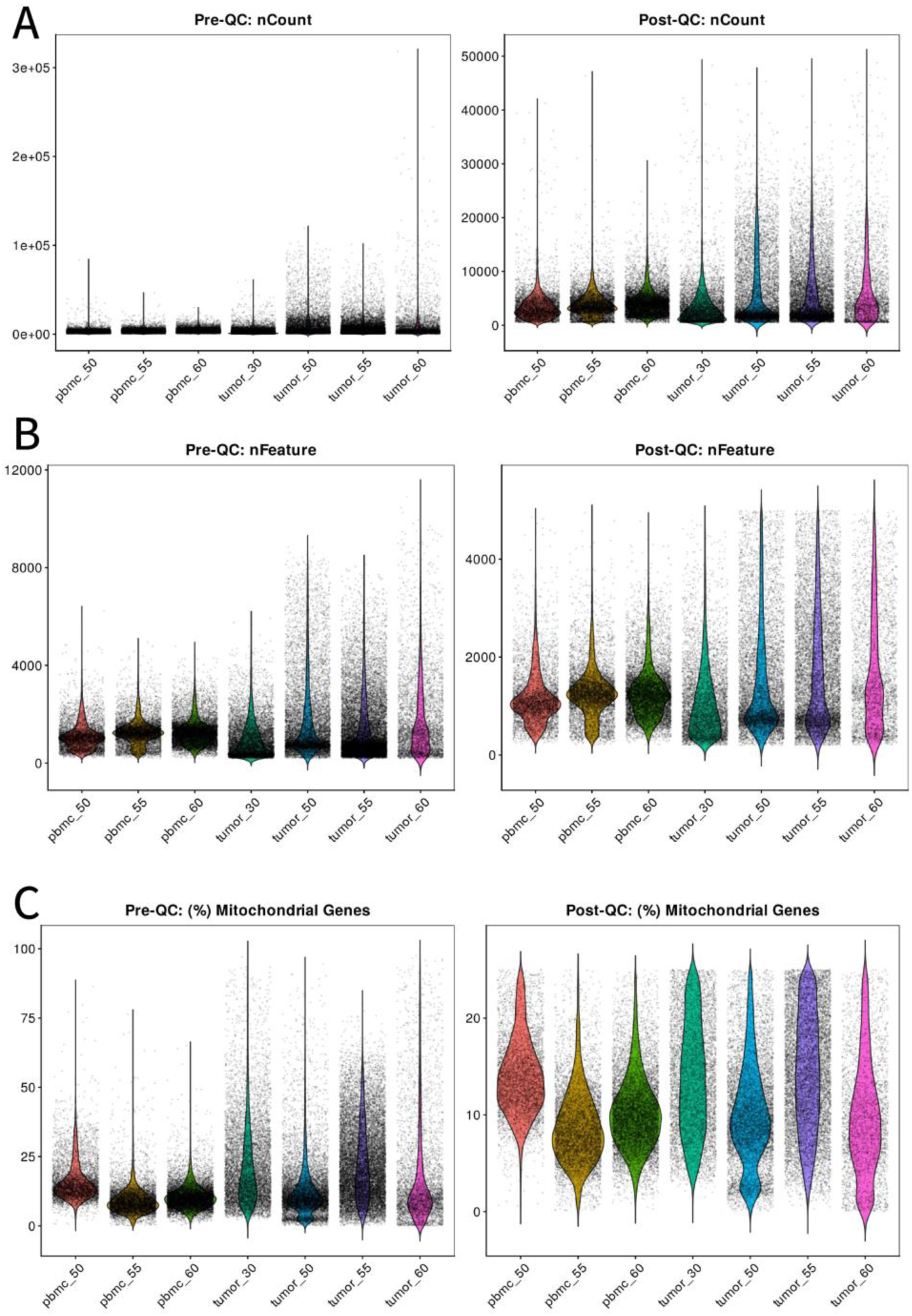
Quality control of GSE217845. Violin plots of (A) total UMI counts per cell (nCount), (B) number of detected genes per cell (nFeature), and (C) percentage of mitochondrial transcripts per cell for each sample before (left) and after (right) quality control. Samples include PBMC and tumor. Filtering thresholds are described in Methods.

**Supplementary Figure S4.**
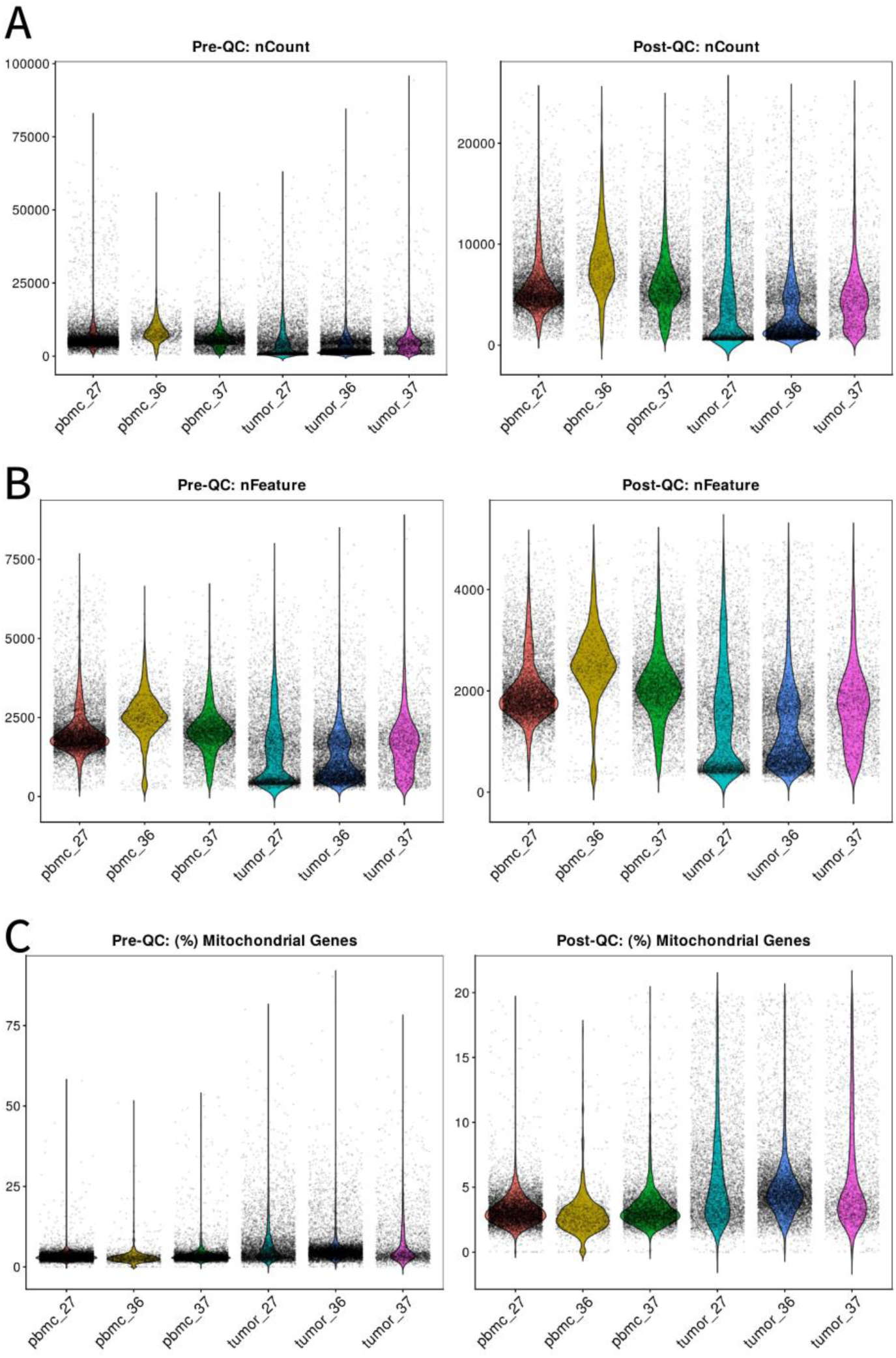
Quality control of GSE300304. Violin plots of (A) total UMI counts per cell (nCount), (B) number of detected genes per cell (nFeature), and (C) percentage of mitochondrial transcripts per cell for each sample before (left) and after (right) quality control. Samples include PBMC and tumor. Cells with fewer than 200 or more than 5,000 detected genes, more than 25,000 UMIs, or more than 20% mitochondrial transcripts were removed.

**Supplementary Figure S5.**
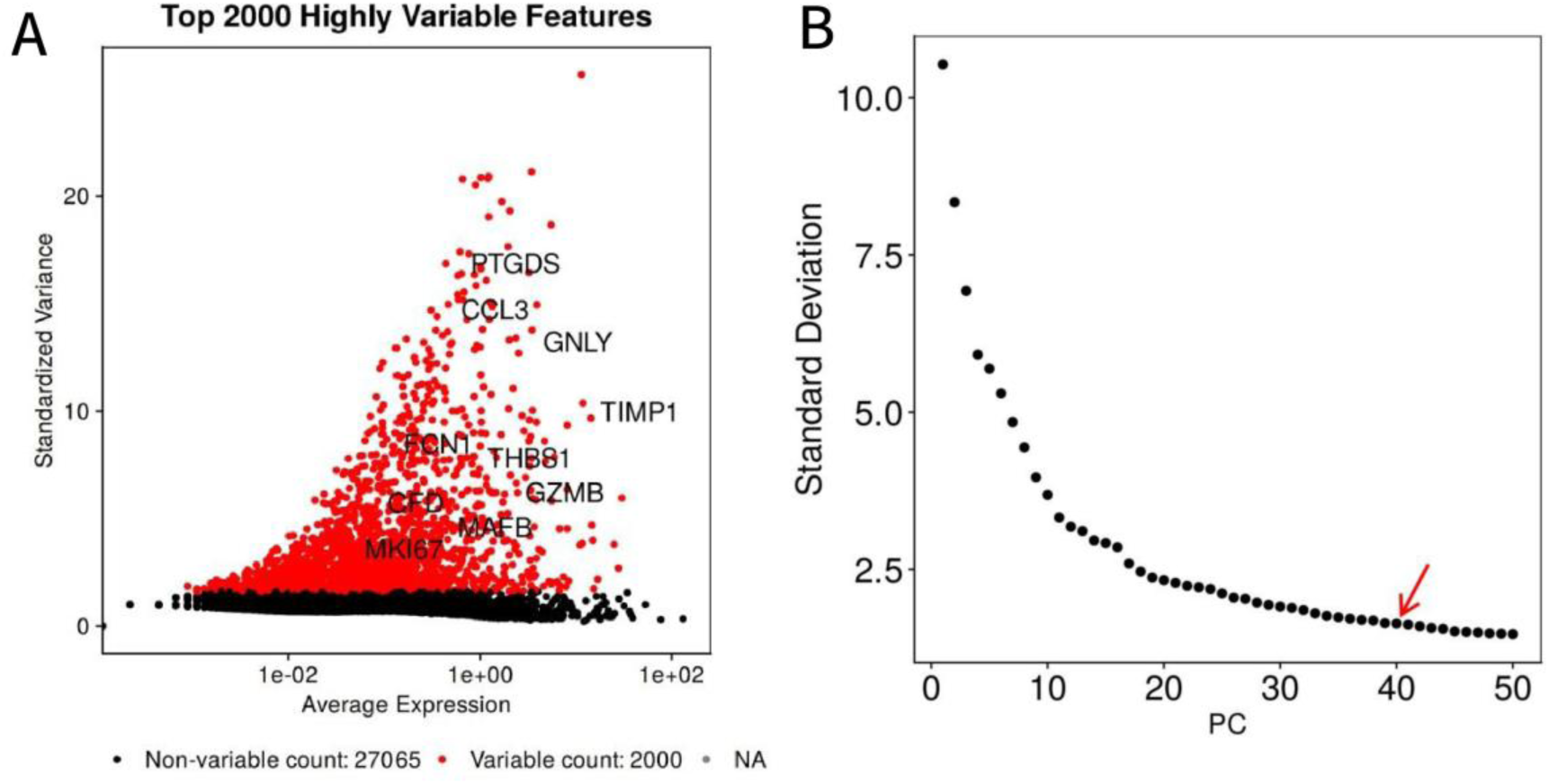
Feature selection and dimensionality reduction. (A) Standardized variance versus average expression for all genes; the top 2,000 highly variable genes are shown in red and selected examples are labeled. (B) Elbow plot of the standard deviation explained by the first 50 principal components; the arrow marks the 40 components retained for downstream analysis.

**Supplementary Figure S6.**
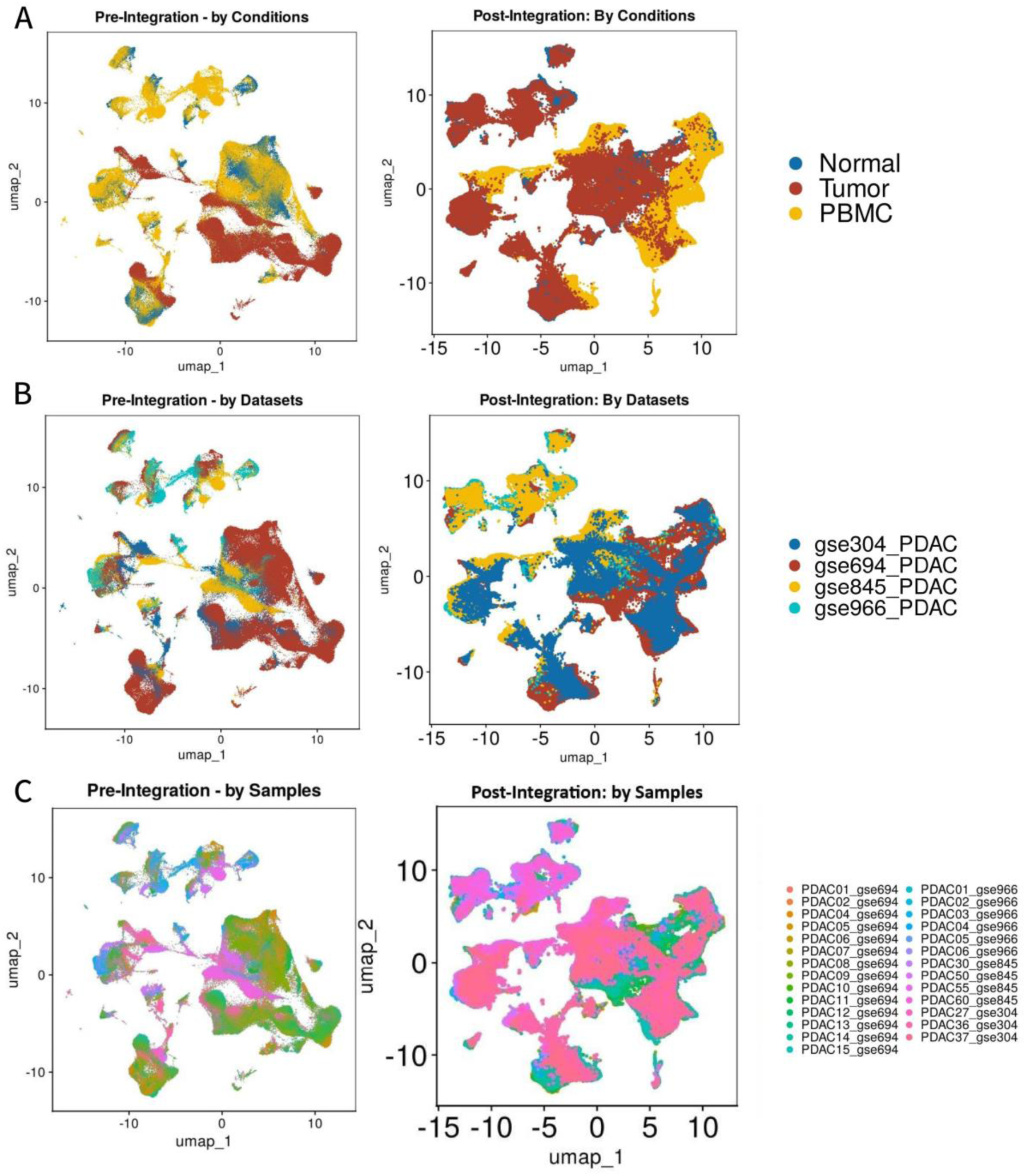
Harmony integration of the four scRNA-seq datasets. UMAP embeddings before (left) and after (right) integration, colored by (A) tissue condition (normal, tumor, and PBMC), (B) source dataset, and (C) sample.

**Supplementary Figure S7.**
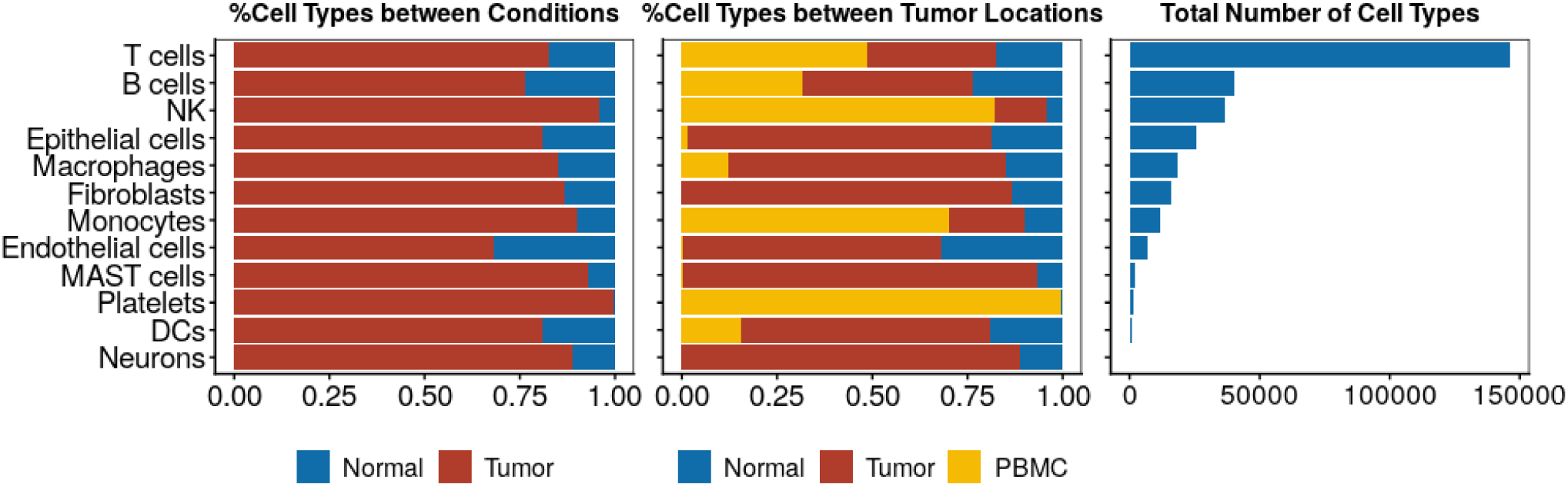
Tissue source and abundance of annotated cell types in the integrated scRNA-seq atlas. (Left) Proportion of cells of each type derived from normal versus tumor samples. (Middle) Proportion of cells of each type derived from adjacent normal tissue, tumor tissue, and peripheral blood mononuclear cells (PBMC). (Right) Total number of cells per annotated cell type across the integrated dataset (324,662 cells from 26 patients).

**Supplementary Figure S8.**
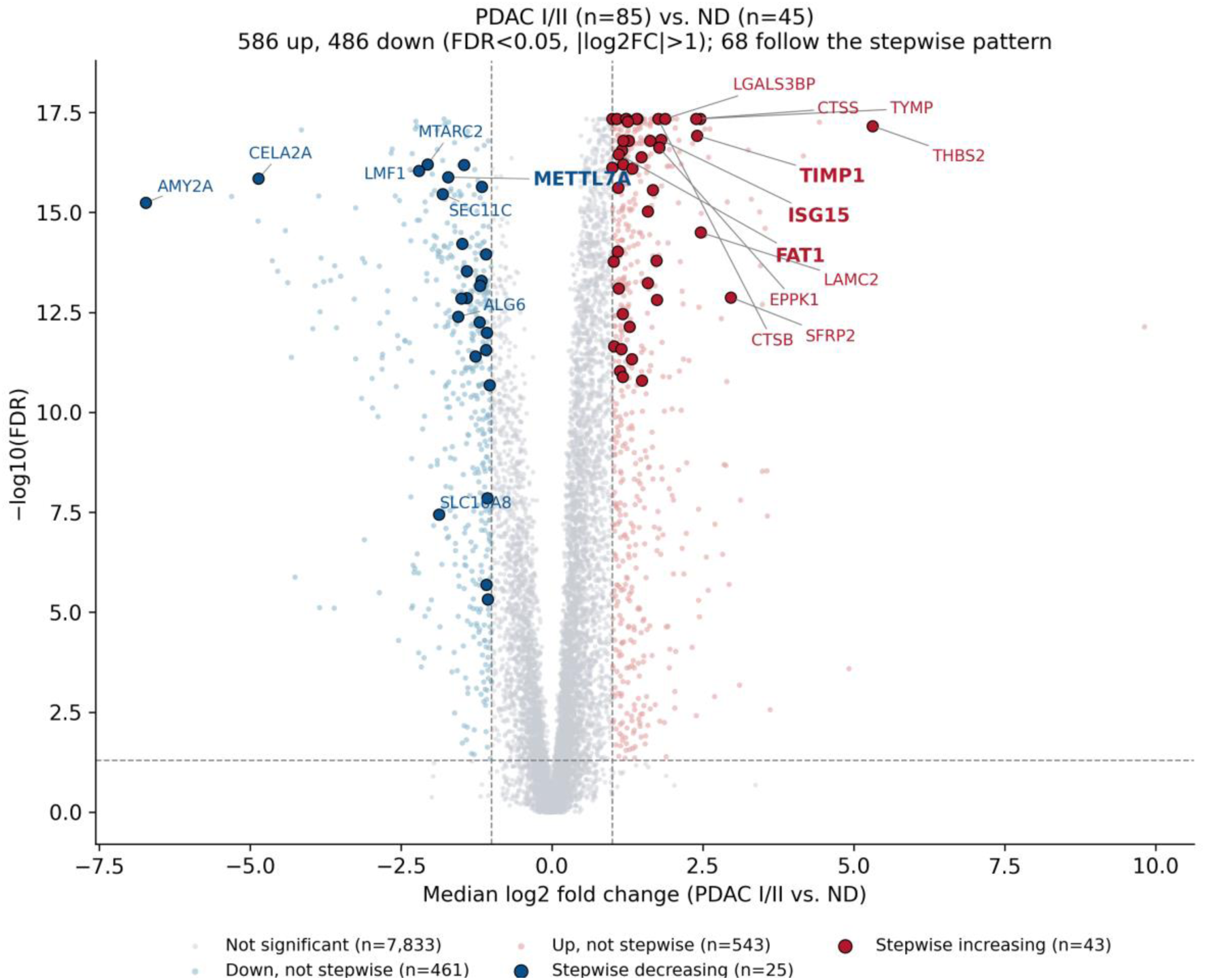
Differential protein abundance between PDAC stage I/II and non-diseased pancreas. Volcano plot of median log2 fold change (PDAC I/II, n = 85, versus ND, n = 45) against −log10(FDR) for 8,905 quantified proteins (two-sided Wilcoxon rank-sum test with Benjamini–Hochberg correction). Dashed lines mark FDR = 0.05 and |log2FC| = 1. Of the 1,072 significant proteins (586 increased, 486 decreased), the 68 that followed the stepwise pattern across the six reference groups are outlined (dark red, increasing, n = 43; dark blue, decreasing, n = 25). Bold labels mark proteins included in candidate panels (TIMP1, ISG15, FAT1, and METTL7A).

**Supplementary Figure S9.**
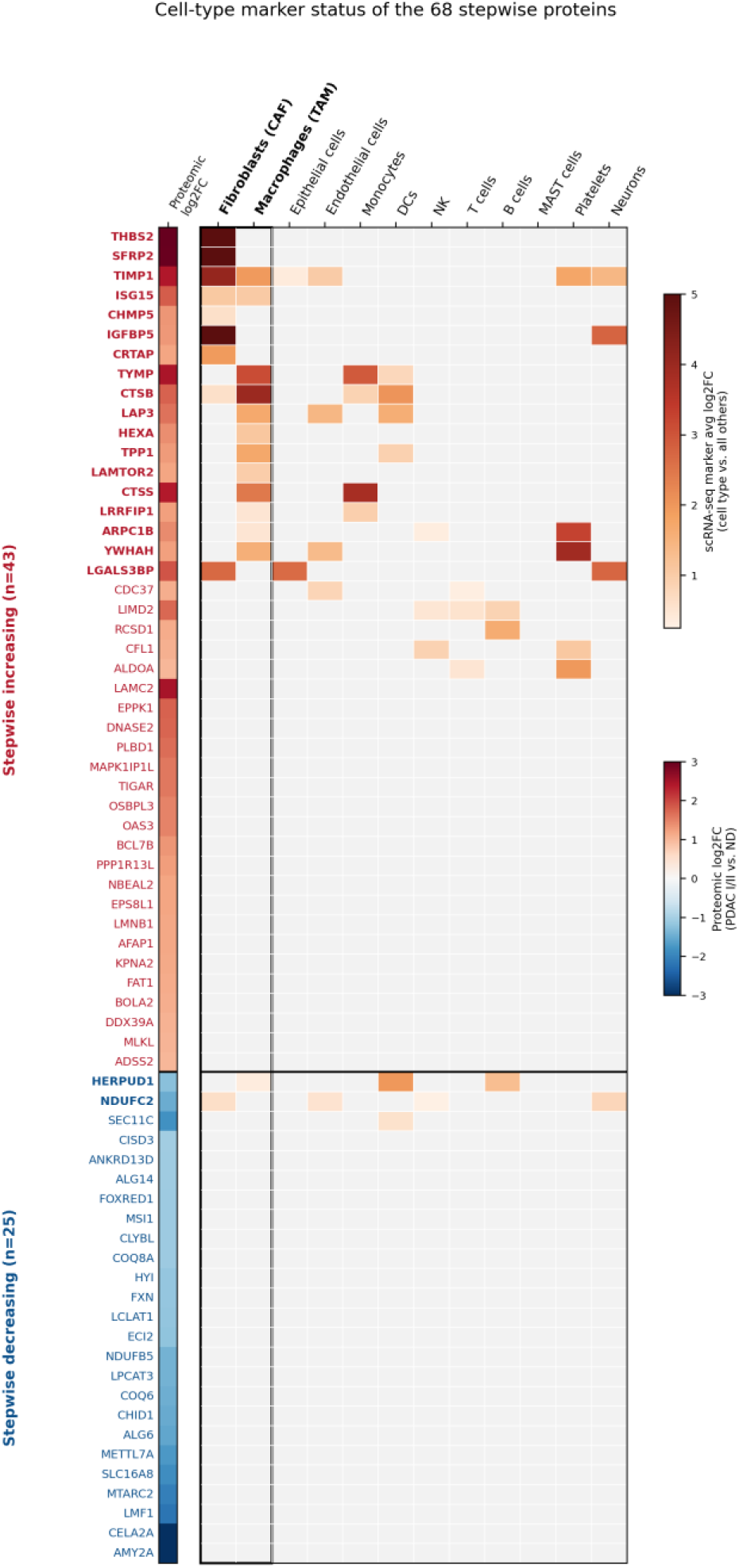
Cell-type marker status of the 68 stepwise proteins. Rows are stepwise-increasing (top, n = 43) and stepwise-decreasing (bottom, n = 25) proteins labeled by gene symbol; the left strip shows proteomic log2 fold change (PDAC I/II versus ND). Heatmap values are the average log2 fold change of each gene in the indicated cell type relative to all other cells (Wilcoxon rank-sum test, Bonferroni-adjusted P < 0.05); grey indicates not a significant marker. Fibroblast (CAF) and macrophage (TAM) columns are boxed, and bold labels mark proteins that are fibroblast or macrophage markers.

**Supplementary Figure S10.**
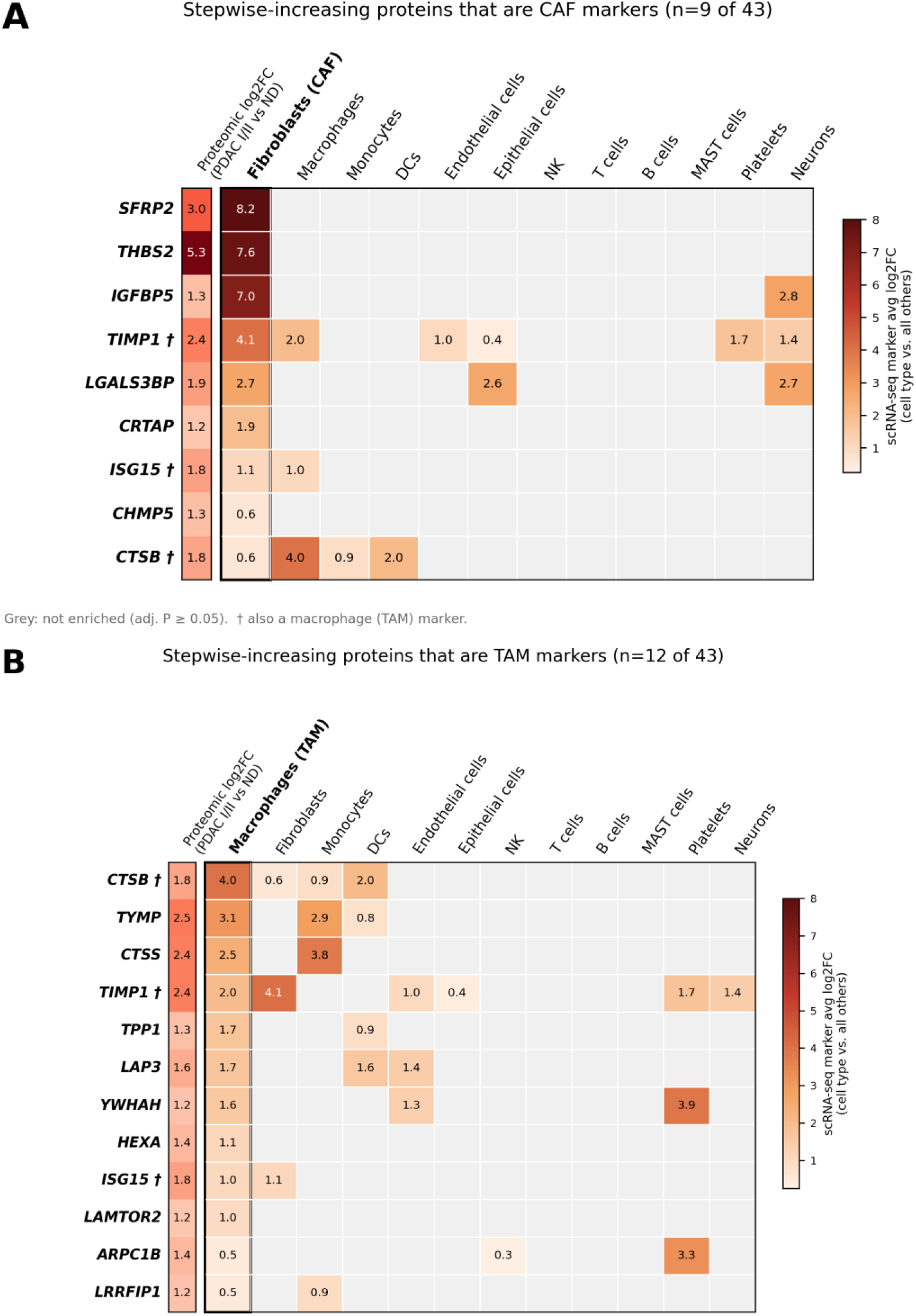
Stepwise-increasing proteins associated with cancer-associated fibroblasts and tumor-associated macrophages. (A) The 9 stepwise-increasing proteins whose genes are fibroblast (CAF) markers and (B) the 12 whose genes are macrophage (TAM) markers, ordered by enrichment in the respective cell type. Values are the scRNA-seq average log2 fold change in each cell type relative to all other cells (Bonferroni-adjusted P < 0.05); grey indicates not a significant marker. The left strip shows proteomic log2 fold change (PDAC I/II versus ND). † Marker of both fibroblasts and macrophages (TIMP1, ISG15, and CTSB).

